# Common and rare variant analyses combined with single-cell multiomics reveal cell-type-specific molecular mechanisms of COVID-19 severity

**DOI:** 10.1101/2021.06.15.21258703

**Authors:** Sai Zhang, Johnathan Cooper-Knock, Annika K. Weimer, Calum Harvey, Thomas H. Julian, Cheng Wang, Jingjing Li, Simone Furini, Elisa Frullanti, Francesca Fava, Alessandra Renieri, Cuiping Pan, Jina Song, Paul Billing-Ross, Peng Gao, Xiaotao Shen, Ilia Sarah Timpanaro, Kevin P. Kenna, VA Million Veteran Program, GEN-COVID Network, Mark M. Davis, Philip S. Tsao, Michael P. Snyder

## Abstract

The determinants of severe COVID-19 in non-elderly adults are poorly understood, which limits opportunities for early intervention and treatment. Here we present novel machine learning frameworks for identifying common and rare disease-associated genetic variation, which outperform conventional approaches. By integrating single-cell multiomics profiling of human lungs to link genetic signals to cell-type-specific functions, we have discovered and validated over 1,000 risk genes underlying severe COVID-19 across 19 cell types. Identified risk genes are overexpressed in healthy lungs but relatively downregulated in severely diseased lungs. Genetic risk for severe COVID-19, within both common and rare variants, is particularly enriched in natural killer (NK) cells, which places these immune cells upstream in the pathogenesis of severe disease. Mendelian randomization indicates that failed NKG2D-mediated activation of NK cells leads to critical illness. Network analysis further links multiple pathways associated with NK cell activation, including type-I-interferon-mediated signalling, to severe COVID-19. Our rare variant model, PULSE, enables sensitive prediction of severe disease in non-elderly patients based on whole-exome sequencing; individualized predictions are accurate independent of age and sex, and are consistent across multiple populations and cohorts. Risk stratification based on exome sequencing has the potential to facilitate post-exposure prophylaxis in at-risk individuals, potentially based around augmentation of NK cell function. Overall, our study characterizes a comprehensive genetic landscape of COVID-19 severity and provides novel insights into the molecular mechanisms of severe disease, leading to new therapeutic targets and sensitive detection of at-risk individuals.

## INTRODUCTION

Infection with severe acute respiratory syndrome coronavirus 2 (SARS-CoV-2) giving rise to coronavirus disease 2019 (COVID-19) has caused a global pandemic with almost unprecedented morbidity and mortality^1^. The severity of COVID-19 is markedly variable ranging from an asymptomatic infection to fatal multiorgan failure. Severity correlates with age and comorbidities^2^ but not exclusively^3^. Indeed, host genetics has been thought to be an essential determinant of severity^4^, but this is poorly understood. Improved tools to identify individuals at risk of severe COVID-19 could facilitate life-saving precision medicine.

There have been several efforts to address the genetic basis of COVID-19 severity^5,6^, including large-scale genome-wide association studies (GWASs)^7,8^ and rare variant approaches^9–12^. However, the biological interpretation of those identified loci has been difficult, partially because of the confounding effects of patient age and comorbidities^13^. The development of novel therapies is likely to result from understanding and modifying the host immune response to the SARS-CoV-2 virus, independent of immutable factors such as age, sex, and general health.

A primary cause of morbidity and mortality in COVID-19 is respiratory disease and specifically, a hyperinflammatory response within the lung that occurs in an age-independent manner^14^. This is the basis of a number of interventions based on immunosuppression^15^, which have repurposed treatments used for other diseases, particularly autoimmune diseases. Efficacy and the side effect profile is likely to be improved by a COVID-19-specific immunomodulatory approach.

Profiles of the immune response associated with severe COVID-19 have produced a number of conflicting observations. These studies have variously linked COVID-19 severity to CD8 T cells^16^, CD19 B cells^17^, eosinophils^18^, and myeloid cells^19^. Single-cell omic profiling has demonstrated the differential function of various immune cell types in severe disease as opposed to mild disease or non-infected condition^20–25^. However, these studies have focused on transcriptomics rather than the underlying genomics, and have been observational rather than predictive.

Failure of the type I interferon response is linked to the incidence of severe COVID-19. SARS-CoV-2 can initially inhibit the normal type I interferon response^26^ in order to facilitate viral replication. This delay is thought to be an essential determinant of a later hyperinflammatory response and consequently of COVID-19 severity^27^. Natural killer (NK) cells form a crucial component of the innate immune response to viral infections. Interestingly, NK cells are activated via the type I interferon response. Genetic evidence suggests that NK cell function is a key determinant of severe COVID-19, including loss-of-function (LoF) variants within an essential NK cell activating receptor, NKG2C, in patients suffering severe COVID-19^28^. A recent study of autoantibodies supports this conclusion by showing that the impaired activation of NK cells, via the type I interferon response in particular, is associated with severe COVID-19^29^. All of this evidence is suggestive of a role for NK cells in severe COVID-19 but not conclusive.

To understand the genetic basis of COVID-19 severity as well as gain insights into its molecular mechanisms, we sought out to integrate the genetic architecture of severe COVID-19, profiled in an age-independent manner, with single-cell-resolution functional profiling of lung tissue. We developed two machine learning frameworks, RefMap and PULSE, for common and rare variant analysis respectively, with increased discovery power compared to traditional methodology. Using our approaches, we identified over 1,000 genes associated with critical illness across 19 cell types, and the cell-type-specific molecular mechanisms underlying severe disease were uncovered. Notably, both common and rare variant analyses underscored the importance of NK cells in determining COVID-19 severity, which extends previous literature^30^. We have developed a prediction model for severe COVID-19 using rare variants profiled by exome sequencing, which achieves sensitive and age- and population-independent risk prediction across multiple cohorts. This prediction method could be particularly useful for targeting medical interventions for individuals where SARS-CoV-2 vaccination is not possible or is not effective^31^. Altogether, our study unveils a holistic genetic landscape of COVID-19 severity and provides a better understanding of the disease pathogenesis, implicating new prevention strategies and therapeutic targets.

## RESULTS

### RefMap analysis of common variants uncovers cell-type-specific genetic basis of COVID-19 severity

We used the RefMap machine learning model (**Methods**) to identify the genomic regions and genes associated with severe COVID-19. Briefly, RefMap is a Bayesian network that combines genetic signals (e.g., allele *Z*-scores) with functional genomic profiling (e.g., ATAC-seq and ChIP-seq) to fine-map risk regions for complex diseases. With RefMap, we can scan the genome for functional regions in which disease-associated genetic variation is significantly shifted from the null distribution. The power of the RefMap model for gene discovery and recovery of missing heritability has been demonstrated in our recent work^32^. Here, to achieve cell-type-specific resolution within multicellular tissue, we modified RefMap to integrate single-cell multiomic profiling of human lungs with COVID-19 GWAS data (**Fig. 1a**). In particular, we obtained summary statistics (COVID-19 Host Genetics Initiative, Release 5, phenotype definition A2; 5,101 cases versus 1,383,241 population controls) from the largest GWAS study of COVID-19^7^, where age, sex, and 20 first principal components were included in the analysis as covariates. Severe COVID-19 was defined by the requirement for respiratory support or death attributed to COVID-19. Human lung single-cell multiomic profiling, including snRNA-seq and snATAC-seq, was retrieved from a recent study of healthy individuals^33^. There are 19 cell types identified in both snATAC-seq and snRNA-seq profiles, including epithelial (alveolar type 1 (AT1), alveolar type 2 (AT2), club, ciliated, basal, and pulmonary neuroendocrine (PNEC)), mesenchymal (myofibroblast, pericyte, matrix fibroblast 1 (matrix fib. 1), and matrix fibroblast 2 (matrix fib. 2)), endothelial (arterial, lymphatic, capillary 1 (cap1), and capillary 2 (cap2)), and hematopoietic (macrophage, B-cell, T-cell, NK cell, and enucleated erythrocyte) cell types. We adopted these 19 cell types as the reference set within lung tissue throughout our study. Based on snATAC-seq peaks called in one or more of the 19 cell types to annotate functional regions, we used RefMap to identify disease-associated genomic regions from the COVID-19 GWAS data, which resulted in 6,662 1kb regions passing the 5% significance threshold (*Q*^+/-^-score>0.95, **Methods**; referred to as RefMap COVID-19 regions). These identified regions were further intersected with open chromatin in individual cell types based on corresponding snATAC-seq peaks, resulting in cell-type-specific RefMap regions (mean per cell type =1732.8, standard deviation (SD)=623.5; **Fig. 1b, Supplementary Table 1**). After removing RefMap regions present in more than one cell type (mean per cell type =121.2, SD=142.7), we observed only a weak correlation between the number of unique RefMap regions and the number of snATAC-seq peaks detected per cell type (Spearman *ρ*=0.40, *P*>0.05; **Fig. 1b**), indicating enrichment of genetic signals within certain cell types.

**Figure 1.**
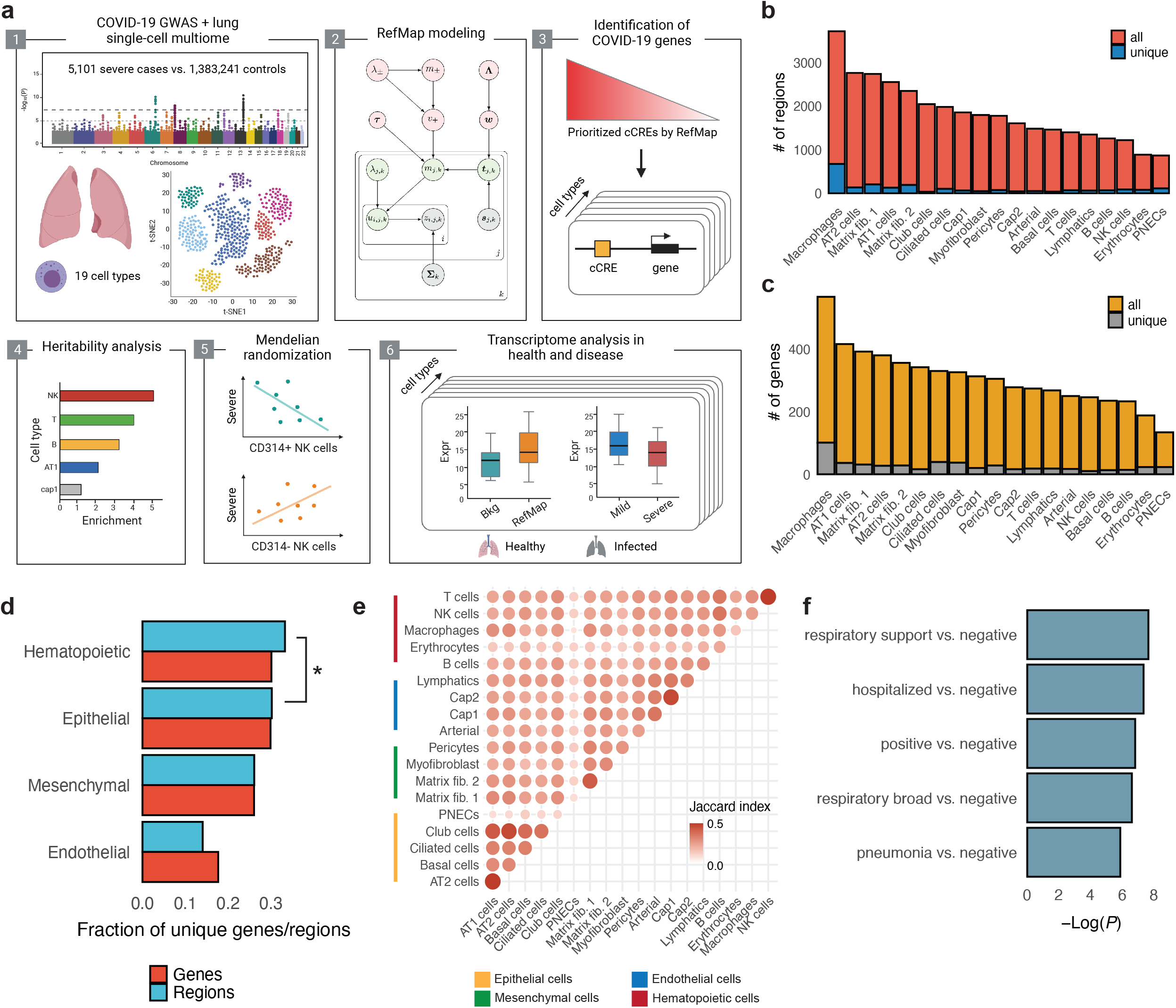
Common variant analysis of COVID-19 severity integrated with lung single-cell multiomics. **a**, Schematic of the study design for fine-mapping cell-type-specific genes from COVID-19 GWAS (Panel 1). The diagram of the RefMap model is shown in Panel 2, where grey nodes represent observations, green nodes are local hidden variables, and pink nodes indicate global hidden variables (**Methods**). Cell-type-specific RefMap genes are mapped using single-cell multiomic profiling (Panel 3). Heritability (Panel 4), Mendelian randomization (Panel 5), and transcriptome analysis (Panel 6) validate the functional importance of RefMap genes, particularly for NK cells, in severe COVID-19. **b**, Total number and number of unique genomic regions containing genetic variation associated with severe COVID-19 for different cell types. **c**, Total number and number of unique genes implicated by genetic variation associated with severe COVID-19 for different cell types. **d**, Fraction of unique genomic regions and genes associated with severe COVID-19 for major cell types. **e**, Similarity between different cell types quantified by the overlap of RefMap genes. Gene set overlapping was calculated by the Jaccard index. **f**, RefMap regions overlap significantly with COVID-19-associated genetic variation in an independent COVID-19 GWAS study. cCRE: candidate cis-regulatory element. *: *P*<0.05.

Next, we sought to map the target genes of RefMap COVID-19 regions in a cell-type-specific manner. In particular, we identified the closest genes that are expressed in the corresponding cell type for individual RefMap regions (**Methods**). In total, we discovered 1,370 genes (referred to RefMap COVID-19 genes; mean per cell type =279.9 and SD=80.3; **Fig. 1c, Supplementary Table 1**) associated with the severe disease. Interestingly, hematopoietic cells have the largest number of unique RefMap regions and genes among all major cell types (**Fig. 1d**); for example, there is a significant enrichment of unique RefMap regions observed for hematopoietic cells versus epithelial cells (*P*=5.2e-03, odds ratio (OR)=1.15, Fisher’s exact test; **Fig. 1d**). This indicates a critical role of immune cells, which are primarily hematopoietic, in the development of severe COVID-19^16–19^. To profile the cell-cell interactions underlying severe COVID-19 from a genetic perspective, we constructed a cell correlation matrix based on the overlap of RefMap genes between cell types (**Fig. 1e**). We discovered that the correlation is strongest between functionally related cells, demonstrating that the RefMap signal is consistent with known biology^33^.

To replicate our findings, we obtained SNPs associated with severe COVID-19 from a GWAS for an entirely independent sample set (the 23andMe cohort, 15,434 COVID-19-positive cases and 1,035,598 population controls)^5^. The total union of RefMap regions is significantly enriched with SNPs associated with multiple COVID-19 phenotypes defined in this new sample set (mean *P*<5e-03, Fisher’s exact test; **Fig. 1f, Supplementary Table 2**; **Methods**). Specifically, the most significant enrichment is with SNPs associated with COVID-19 requiring respiratory support (mean *P*=4.68e-04, Fisher’s exact test; **Fig. 1f**). We further performed the enrichment analysis per cell type; only RefMap regions associated specifically with T cells and NK cells are significantly enriched with disease-associated SNPs across all measured COVID-19 phenotypes (mean *P*<0.05, Fisher’s exact test; **Supplementary Table 2**).

### Heritability analysis and Mendelian randomization link NK cell function to COVID-19 severity

The LD score regression (LDSC)^34^ has been used to measure the total SNP-based heritability (*h*^2^) from the GWAS study of severe COVID-19 (COVID-19 Host Genetics Initiative, Release 5, phenotype definition A2)^7^. Here, we examined the partitioning of SNP-based heritability for severe COVID-19 within RefMap genes (**Methods**). We discovered that the heritability of severe COVID-19 is significantly enriched for RefMap genes (OR=4.6, standard error (SE)=0.78, *P*=1.55e-07; **Fig. 2a, Supplementary Table 3**). We compared the proportion of SNP-based heritability captured by RefMap to other methods (**Methods**), including naïve GWAS^7^ and MAGMA^35^. The proportion of heritability within naïve GWAS genes is 0.15 compared to 0.37 within MAGMA genes, but 0.77 within RefMap genes (**Fig. 2b, Supplementary Table 3**), representing a five-fold improvement in the recovered heritability based on RefMap over traditional methods. The proportion of SNP-based heritability for hospitalized COVID-19 (COVID-19 Host Genetics Initiative, Release 5, phenotype definition B2) within RefMap genes is 0.62 and within COVID-19 independent of severity (COVID-19 Host Genetics Initiative, Release 5, phenotype definition C2) it is 0.52 (**Fig. 2b, Supplementary Table 3**). In both cases the improvement in captured heritability based on RefMap compared to traditional methods is three-fold. Consistent with the design of our model, the recovered heritability is highest in severe COVID-19.

**Figure 2.**
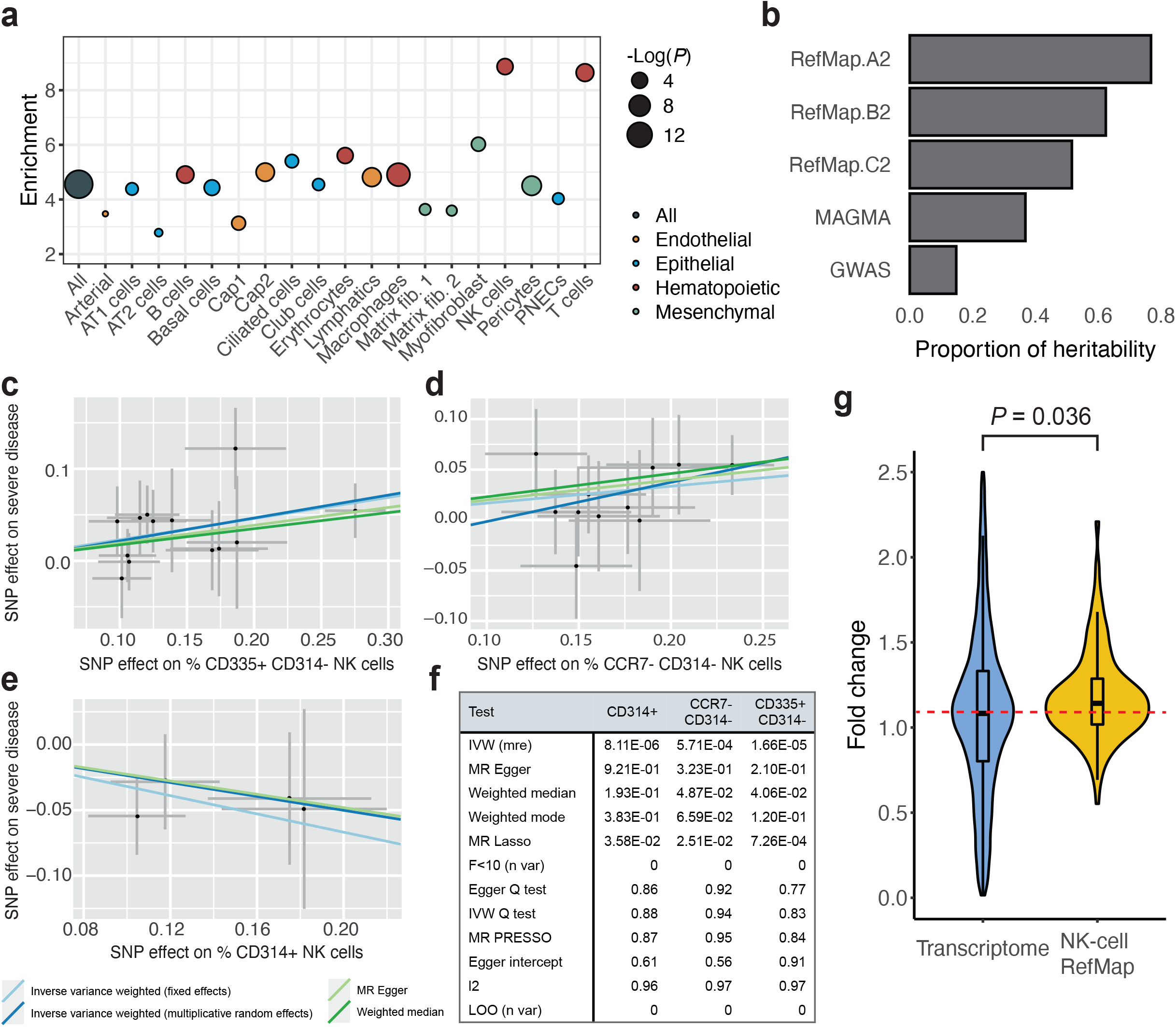
Severe-COVID-19-associated common variants are linked to NK cell function. **a**, Heritability enrichment estimated by LDSC for different cell types. Enrichment was calculated as the proportion of total SNP-based heritability adjusted for SNP number. **b**, Proportion of SNP-based heritability associated with risk genes identified using RefMap or conventional methodology. **c, d, e**, Significant Mendelian randomization results for three exposures linked to severe COVID-19, including blood counts of (**c**) CD335+ CD314-, (**d**) CCR7-CD314-, and (**e**) CD314+ NK cells. **f**, Sensitivity analyses and robust tests for MR analyses (**Methods**). **g**, Comparative gene expression analysis of NK-cell RefMap genes in NKG2D+ and NKG2D-NK cells. Fold change was calculated as the ratio of gene expression levels in NKG2D+ NK cells to NKG2D-NK cells. The transcriptome was defined by all the expressed genes (with at least one UMI (unique molecular identifier)) in NK cells. Violin plots show the distributions of fold change values within each group, and boxplots indicate the median, interquartile range (IQR), Q1−1.5×IQR, and Q3+1.5×IQR. The red dashed line denotes the median value of fold change distribution for the transcriptome.

Next, we used cell-type-specific RefMap genes to determine which cell types are involved in the development of severe COVID-19. Specifically, we calculated the partitioned heritability per cell type within the severe COVID-19 GWAS (A2) and also within GWAS for hospitalized versus non-hospitalized COVID-19 (B2) and COVID-19 versus population (C2) (**Methods**). For severe COVID-19, of all 19 cell types tested, NK cells are the most enriched with SNP-based heritability (OR=8.87, SE=3.68, *P*=0.016; **Fig. 2a, Supplementary Table 3**). The same is also true for hospitalized COVID-19 (OR=10.57, SE=4.95, *P*=0.039), but not for COVID-19 irrespective of severity (OR=5.74, SE=3.09, *P*=0.077; **Supplementary Table 3**). Thus, we conclude that NK cell function is enriched with severe disease-associated genetic variation.

Two-sample Mendelian randomization (MR) facilitates identification of a causal relationship between an exposure and an outcome^36^. We examined whether NK cell populations measured in the blood are causally related to severe COVID-19. In total, 46 GWAS measures of NK cell subtypes were identified^37^ (**Methods**). After harmonizing exposure and outcome genetic instruments, we excluded tests with less than five SNPs (**Methods**). With MR, three exposures were shown to be causally related to severe COVID-19 after correcting for multiple testing (*P*<1e-03, multiplicative random effects (MRE), inverse-variance weighted (IVW)). All three exposures relate to NKG2D/CD314 expression on the cell surface, where a higher number of NKG2D/CD314-cells was linked to severe COVID-19 (A2) (**Figs. 2c-d**) and a higher number of NKG2D/CD314+ cells is protective (**Fig. 2e**). Evidence of genetic pleiotropy (MR PRESSO intercept not significantly different from zero, *P*>0.05; **Fig. 2f**) or instrument heterogeneity (*P*>0.05, Cochran’s *Q* test, and *I*^2^_GX_ >0.95; **Fig. 2f**) are not evident. Moreover, robust measures are significant for all three exposures (**Fig. 2f**). We also tested the identical phenotypes with alternative COVID-19 phenotype GWAS; we discovered that CD335+ CD314-cell counts are also causally associated with hospitalized COVID-19 (B2), and with COVID-19 independent of severity (C2) (**Supplementary Fig. 1**), but in each case the effect size is reduced compared to severe COVID-19 (A2). NKG2D/CD314 is a primary receptor responsible for NK cell activation^38^ and in light of this, we conclude that severe COVID-19 is associated with a loss of NK cell cytotoxicity rather than a gain of function linked to NKG2D/CD314-cells.

Inspired by our MR analysis, we further tested if the expression of RefMap genes reflects a functional difference between NKG2D/CD314+ and NKG2D/CD314-cells. We examined the expression levels of NK-cell RefMap genes based on scRNA-seq data from healthy lungs^39^, and discovered that RefMap gene expression is higher within NKG2D/CD314+ cells than NKG2D/CD314-cells (*P*=0.036, one-tailed Wilcoxon rank-sum test; **Fig. 2g**). This further supports the functional significance of RefMap genes in COVID-19 severity and associates the genetic risk of severe COVID-19 directly with NK cell activity.

### Transcriptome analysis supports the functional significance of RefMap genes in health and severe COVID-19

To link RefMap COVID-19 genes to the underlying biology, we first performed functional enrichment analyses based on gene ontology (GO) and KEGG pathways^40^ (**Figs. 3a** and **3b, Supplementary Tables 4** and **5**). We observed that RefMap NK-cell genes are enriched with pathways and ontologies related to intra- and intercellular signalling important for NK cell activation, including “Phospholipase D signalling pathway”^41^, “Antigen processing and presentation”, “regulation of small GTPase mediated signal transduction (GO:0051056)”^42^, and “regulation of intracellular signal transduction (GO:1902531)” (adjusted *P*<0.1; **Figs. 3a** and **3b, Supplementary Tables 4** and **5**). This is consistent with the hypothesis that COVID-19 severity is determined by failed activation of NK cells. Furthermore, the pathway with the highest enrichment is “human immunodeficiency virus (HIV) 1 infection” (adjusted *P*=3e-04; **Fig. 3b**). Since HIV-1 works to suppress NK cell activation^43^, and NK cell function has been associated with an effective immune response to HIV^44^, this result is also consistent with a role of NK cells in severe COVID-19. Other cell-type-specific RefMap gene lists are also enriched with relevant biological pathways. For example, AT2-cell genes are linked to pathways associated with viral infection such as ‘human papillomavirus infection’ and ‘viral carcinogenesis’ (adjusted *P*<0.1; **Supplementary Table 5**), which is consistent with the established role of AT2 cells as the initial site of SARS-CoV-2 entry into host cells^45^. T-cell genes are enriched with ‘IL17-signalling pathway’ (adjusted *P*=0.021; **Supplementary Table 5**) which is interesting in light of previous literature highlighting the production of IL-17 by T cells from COVID-19 patients as a potential therapeutic target^46^.

**Figure 3.**
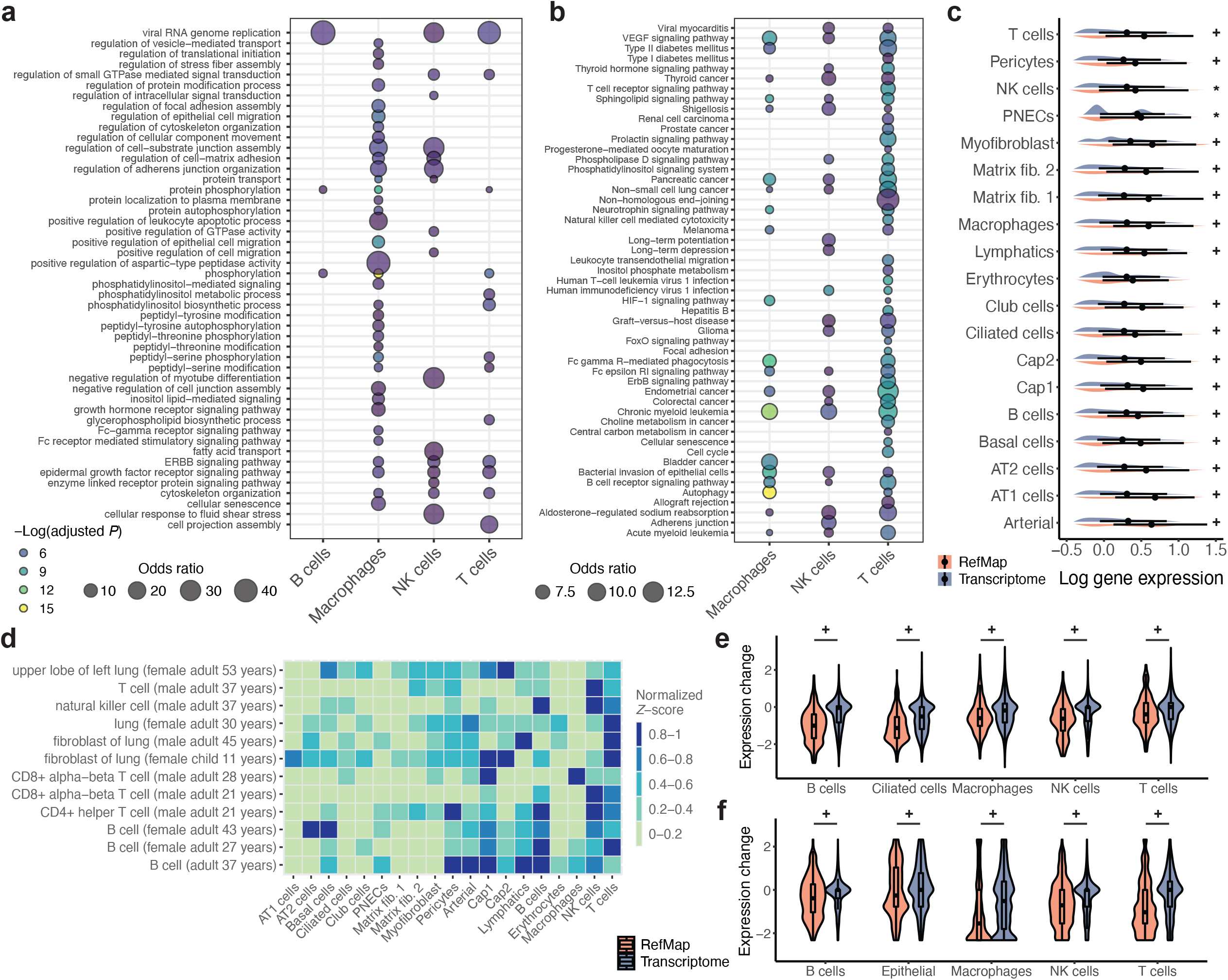
Functional enrichment and transcriptome analyses of RefMap COVID-19 genes. **a**, Gene Ontology (GO) terms that are significantly enriched in cell-type-specific RefMap gene lists corresponding to hematopoietic cell types; only terms with adjusted *P*<0.05, OR>3, and character number<60 are visualized. **b**, KEGG Pathways that are significantly enriched in cell-type-specific RefMap gene lists corresponding to hematopoietic cell types; only terms with adjusted *P*<0.05, OR>5, and character number<50 are visualized. **c**, Gene expression analysis of RefMap genes across different cell types in healthy lungs. The transcriptome was defined as the total set of expressed genes for each cell type (**Methods**). Violin plots show the distributions of log expression levels within each group, and point plots indicate the median and IQR. **d**, Overlap between cell-type-specific RefMap regions and H3K27ac and H3K4me3 ChIP-seq peaks from ENCODE lung and immune cell samples. *Z*-scores calculated by regionR^49^ (1,000 permutations) were normalized into the 0-1 range for visualization. **e, f**, Comparative gene expression analysis of cell-type-specific RefMap genes in severe COVID-19 patients versus moderately affected patients based on scRNA-seq datasets from (**e**) Ren et al. and (**f**) Liao et al., respectively. The *Z*-score of Wilcoxon rank-sum test was used to indicate the gene expression change between severe and moderate patient groups, where a positive value means higher gene expression in severe patients. The Benjamini-Hochberg (BH) procedure was used to calculate FDRs throughout the study. Violin plots show the distribution of gene expression changes within each group, and boxplots indicate the median, IQR, Q1−1.5×IQR, and Q3+1.5×IQR. *: FDR<0.1. +: FDR<0.01.

Next, we investigated the baseline expression pattern of RefMap genes in healthy lungs. In particular, we calculated mean expression levels of genes in different cell types based on the lung snRNA-seq data from Wang et al.^33^, and then compared the expression of RefMap genes with the total set of expressed genes in each cell type. Interestingly, although the gene expression level was not an input to the RefMap model, RefMap genes are expressed at a higher level compared to the background transcriptome in all 19 cell types, including immune and epithelial cells (false discovery rate (FDR)<0.1, one-tailed Wilcoxon rank-sum test; **Fig. 3c**) with the exception of pericytes (FDR=0.11, *Z*-score=1.25); it is interesting to note that pericytes may be downstream in the pathogenesis of COVID-19 because they are protected by an endothelial barrier^47^. This supports the functional significance of RefMap genes across multiple cell types in healthy human lungs. As a negative control, we performed a similar expression comparison between non-developmental genes and all expressed genes in lungs, which yielded no significant difference (**Supplementary Fig. 2**; **Methods**). In summary, our transcriptome analyses indicate that RefMap genes are expressed above background in relevant cell types, supporting their important role in lung function.

To obtain further insights into the function of RefMap COVID-19 regions, we tested whether RefMap regions are enriched with cell-type-specific candidate cis-regulatory elements (cCREs, or enhancers and promoters) defined by H3K27ac and H3K4me3. We obtained cCREs for lung tissues and primary cells from the ENCODE project^48^ (**Supplementary Table 6**), and examined the overlap between those cCREs and RefMap regions by permutation test^49^. We discovered that RefMap regions specific to immune cells (e.g., T cells, B cells, and NK cells) are significantly enriched with cCREs in corresponding cell types (FDR<0.1; **Fig. 3d**). For other cell types, RefMap regions are generally enriched with cCREs from lung tissue (FDR<0.1; **Fig. 3d**). These observations are consistent with an important role of RefMap regions in the regulation of gene expression. Moreover, the enrichment with cCREs across a variety of individuals and datasets supports the generalizability of the genetic architecture defined by RefMap. A similar association between genome-wide snATAC-seq peaks and cCREs was also observed (**Supplementary Fig. 3**).

We have shown that RefMap COVID-19 regions are enriched within promoters and enhancers responsible for regulating gene expression in healthy lung tissue. On this basis, we hypothesized that genetic variation within RefMap regions would alter the expression of corresponding target genes in the context of severe disease. Specifically, we proposed that RefMap genes would be expressed at a lower level in lung tissue from severe COVID-19 patients than moderately affected patients. To validate this hypothesis, we obtained scRNA-seq data from the respiratory system for a large COVID-19 cohort^23^, including 12 bronchoalveolar lavage fluid (BALF) samples, 22 sputum samples, and 1 sample of pleural fluid mononuclear cells (PFMCs) from 27 severely and 8 mildly affected patients. Severity was classified based on the World Health Organization (WHO) guidelines (https://www.who.int/publications/i/item/WHO-2019-nCoV-clinical-2021-1). For individual cell types, we compared the expression level of RefMap genes in severe patients versus moderately affected patients (**Methods**). Compared to the background transcriptome, we observed that RefMap genes are relatively lower expressed in severe patients in corresponding cell types than in moderate patients (FDR<0.01, one-tailed Wilcoxon rank-sum test; **Fig. 3e**), supporting the functional significance of RepMap genes in severe COVID-19. As a replication experiment, we carried out a similar analysis based on an independent COVID-19 scRNA-seq dataset^22^, including 9 BALF samples from 6 severe patients and 3 moderate patients (**Methods**). The lower expression of RefMap genes in severe patients is consistent across multiple cell types (FDR<0.01, one-tailed Wilcoxon rank-sum test; **Fig. 3f**). Altogether, these transcriptome-based orthogonal analyses are consistent with the hypothesis that identified cell-type-specific RefMap genes are functionally linked to COVID-19 severity.

### PULSE analysis of rare variants enables population-independent prediction of COVID-19 severity

RefMap utilizes common genetic variation profiled in GWAS. Biological dysfunction can also be determined by rare variants and therefore we assessed whether there is a significant burden of severe-COVID-19-associated rare variants within RefMap genes. First, we employed a standard methodology using SKAT^50^ rare-variant burden testing applied to whole-exome sequencing (WES) data from the GEN-COVID cohort^51^, including non-elderly patients who suffered severe COVID-19 requiring respiratory support, and individuals who suffered non-severe COVID-19 not requiring hospitalization (**Methods**). No individual gene is enriched with significant genetic burden after adjusting for multiple testing (**Supplementary Fig. 4**). This is true whether we tested genome-wide or only for RefMap COVID-19 genes. However, for one subset of cell-type-specific RefMap genes, the median *P*-value was lower than expected: NK cells (*P*<0.05, permutation test; **Fig. 4a**; **Methods**). This result from the analysis of rare genetic variation in an independent cohort is convergent with our common variant analysis, highlighting NK cell biology as a critical determinant of COVID-19 severity.

**Figure 4.**
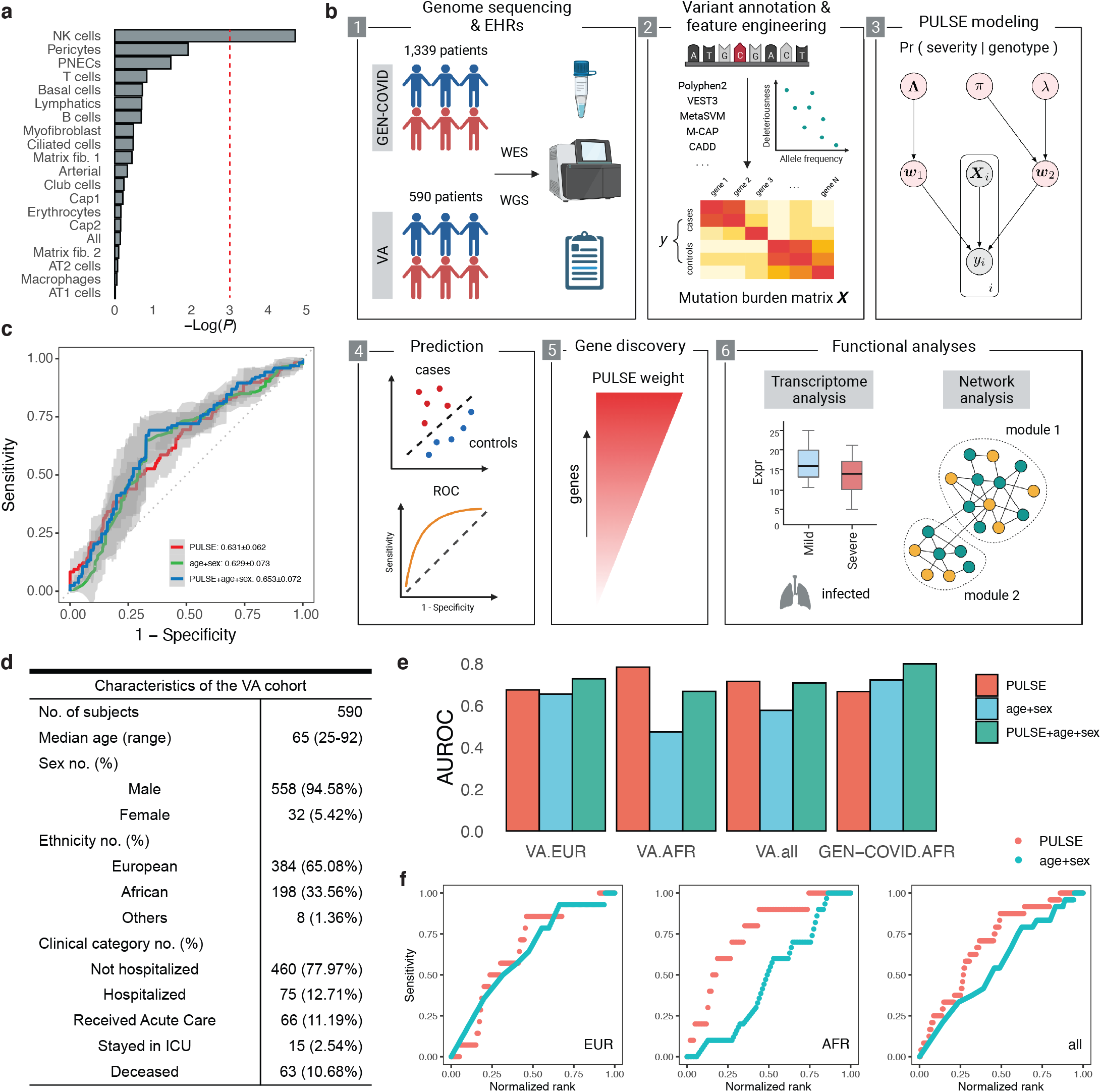
Rare variant analysis informs individual risk of critical illness of COVID-19. **a**, Enrichment analysis of cell-type-specific RefMap COVID-19 genes with rare variants using SKAT burden testing. The red dashed line indicates *P*=0.05. **b**, Schematic of the study design for our rare variant analysis based on PULSE. We examine two independent cohorts in which rare variants were profiled by different technologies: whole-exome sequencing (WES) and whole-genome sequencing (WGS) (Panel 1). Variants are annotated using ANNOVAR (Panel 2) and encoded as input for the PULSE model (Panel 3, **Methods**), where grey nodes are observations and pink nodes represent hidden variables. PULSE is trained to differentiate cases and controls (Panel 4), where the gene weights are useful for gene discovery (Panel 5). Functional characterization of risk genes is performed based on scRNA-seq and PPIs (Panel 6). **c**, Receiver operating characteristic (ROC) curves of different models, including PULSE, age+sex, and integrative models, in the 5-fold cross-validation. Solid lines represent the mean values, and the grey area indicates the standard errors. **d**, Summary statistics of the VA COVID-19 cohort. **e**, AUROC (area under the receiver operating characteristics) scores of predictions in multiple test datasets. Prediction performance is shown for PULSE, age+sex, and integrative models. **f**, Comparison of prediction sensitivity between PULSE and age+sex models. EHRs: electronic health records.

Traditional rare-variant burden testing failed to identify any enrichment of COVID-19-associated variants within a single gene although there is significant enrichment in the group of 236 NK-cell RefMap genes. This suggests that traditional burden testing is underpowered when applied to the GEN-COVID dataset. We decided to develop a new method with increased sensitivity. Here we present PULSE (**<u>p</u>**robabilistic b**<u>u</u>**rden ana**<u>l</u>**ysi**<u>s</u>** based on functional **<u>e</u>**stimation), a discriminative Baysian network that integrates functional annotations of rare variants to model the relationship between genotype and phenotype (**Fig. 4b**; **Methods, Supplementary Notes**). In particular, PULSE combines multiple predictions of functional effects for different types of variants, including missense, nonsense, splicing-site, and small insertion-deletion (indel) mutations (**Supplementary Table 7**). After aggregating those functional scores for individual genes, PULSE learns the importance of different annotations and genes from the training data and maps the phenotype from the genotype in a bilinear form (**Fig. 4b**; **Methods**). With PULSE as a discovery-by-prediction strategy, we are able to (i) predict individual phenotypes from personal genotypes and (ii) discover phenotype-associated genes by model interpretation.

We applied PULSE to study rare genetic variants associated with COVID-19 severity based on the GEN-COVID cohort of whole-exome sequencing from 1,339 COVID-19 patients with 5 severity gradings^52^ (**Fig. 4b, Supplementary Table 8**; **Methods**). After quality controls (QCs), we constructed a discovery cohort (training dataset) of non-elderly European (EUR) adults (age >30 and <60 years) who were critically ill (cases, *n*=109) or not hospitalized (controls, *n*=269) (**Methods**). There is no significant age difference between cases and controls after filtration (*P*=0.29, Wilcoxon rank-sum test; **Supplementary Fig. 5**). We then performed genome annotation^53^ and feature engineering (**Methods**), where only rare variants (i.e., absent from the EUR cohort within the 1000 Genomes Project Phase 3^54^) in autosomes were utilized for downstream analysis. To test the prediction performance of PULSE, we first performed 5-fold cross-validation (CV) based on the GEN-COVID discovery cohort, where a mean AUROC (area under the receiver operating characteristics) of 0.631 was achieved (SE=0.062; **Fig. 4c**). This demonstrates the predictability of COVID-19 severity from personal genomes. The AUROC scores (0.629±0.073) of a logistic regression model built from patient age and sex information are comparable to the PULSE genetic model (**Fig. 4c**). However, combining scores (by averaging) of PULSE and age+sex produced a further improvement in prediction performance (AUROC=0.653±0.072; **Fig. 4c**), demonstrating that host genetics is relatively independent of the effect of age and sex on disease severity. We note that since we removed the age bias in the discovery cohort for genetic concentration, the largest contribution in the age+sex model came from the sex information (model coefficients: 1.33±0.052 for sex versus 0.001±0.004 for age; **Supplementary Fig. 6**). We also note that in our PULSE model only autosomal variants were considered to remove the effect of sex in genetic modelling.

To further validate the prediction power of PULSE, we analyzed whole-genome sequencing (WGS) data of an independent cohort from the Veterans Health Administration (VA), consisting of 590 COVID-19 patients with variable disease severity (**Fig. 4d, Supplementary Table 9**; **Methods**). Extensive QCs (without filtering based on ancestry) resulted in 571 genomes (**Methods**). Genome annotation and feature engineering were conducted as for the GEN-COVID cohort. Similarly, to remove the effect of age, we focused on non-elderly adults (age >30 and <65 years) who were critically ill or not hospitalized, yielding 243 samples (24 cases and 219 controls). In this analysis, we relaxed the upper threshold of age from 60 to 65 years to include more samples in testing. The PULSE model trained on the whole GEN-COVID cohort was applied to predict severity within the VA EUR samples (14 cases versus 125 controls). We found that PULSE succeeded in predicting severe disease solely from personal genomes for this independent cohort with an AUROC of 0.675 (**Fig. 4e**).

Next, we asked if the prediction accuracy is generalizable across different populations. We constructed a test set of non-EUR non-elderly adults (age >30 and <60 years) that passed all other QC criteria within the GEN-COVID cohort, resulting in 12 cases (critically ill) and 6 controls (not hospitalized). The PULSE model trained on EUR samples was then applied to this non-EUR dataset, yielding AUROC of 0.667, which is comparable to the prediction solely based on age and sex (AUROC=0.722; **Fig. 4e**). Combining two scores further increased the prediction performance (AUROC=0.799; **Fig. 4e**). Furthermore, we applied the same trained PULSE model to predict severe disease for African (AFR) individuals (10 cases versus 92 controls) within the VA cohort. Similarly, we discovered that PULSE succeeded in the cross-population prediction with an AUROC of 0.784 (**Fig. 4e**). A similar result was observed for the whole VA dataset with mixed populations (AUROC=0.716; **Fig. 4e**). These results demonstrate the prediction power of PULSE and suggest that the rare-variant genetic architecture of COVID-19 severity is conserved across multiple populations. Importantly, we observed that the prediction in the VA cohort based on just age and sex information trained on the GEN-COVID cohort is inferior to PULSE (AUROC=0.655, 0.474, and 0.577 for EUR, AFR, and all samples, respectively; **Fig. 4e**). This may be linked to the different sex distribution with fewer females in the VA cohort (**Fig. 4d, Supplementary Figs. 7** and **8**), but is further evidence of the robustness of host genetic signals in determining COVID-19 severity and demonstrates that the PULSE prediction is independent of age and sex.

We investigated additional performance measures including sensitivity and specificity based on different cutoffs. Importantly, although the specificity scores are comparable between PULSE and age+sex models cross cutoffs (**Supplementary Fig. 9**), we discovered that PULSE yielded a significantly higher sensitivity (median values: 0.857 versus 0.688, 0.900 versus 0.525, and 0.875 versus 0.560 for EUR, AFR, and all VA samples, respectively; **Fig. 4f**). High sensitivity is important for the clinical application of severity prediction to guide the identification of at-risk individuals. Predictions for the GEN-COVID non-EUR samples yielded similar sensitivity and specificity (**Supplementary Fig. 10**).

### Common and rare variant analyses of severe COVID-19 converge on NK cell function

The trained PULSE model assigns a weighting to individual genes as a measure of association between gene function and severe COVID-19 (**Supplementary Fig. 11**; **Methods**), where a larger weight indicates a higher gene mutation burden in severe patients. To test for the convergence between our common and rare variant analyses, we compared the absolute values of model weights for RefMap genes per cell type with all genes considered by the PULSE model. After correcting for multiple testing, we concluded that for all cell types, RefMap genes tend to have weights with larger absolute values, indicating an association with severe COVID-19 (FDR<0.01, one-tailed Wilcoxon rank-sum test; **Fig. 5a**). Common and rare genetic variations are largely independent^55–57^, and therefore, this convergence of common and rare variant signals indicates shared biology underlying severe disease. Among all cell types, club-cell RefMap genes are the most enriched with PULSE genes (FDR<1e-05, *Z*-score=5.33; **Fig. 5a**). Interestingly, of hematopoietic cells, NK cell genes are the most enriched with PULSE genes (FDR=1.1e-03, *Z*-score=3.16; **Fig. 5a**), consistent with our previous conclusion that NK cells are an essential component of the immune response against SARS-CoV-2.

**Figure 5.**
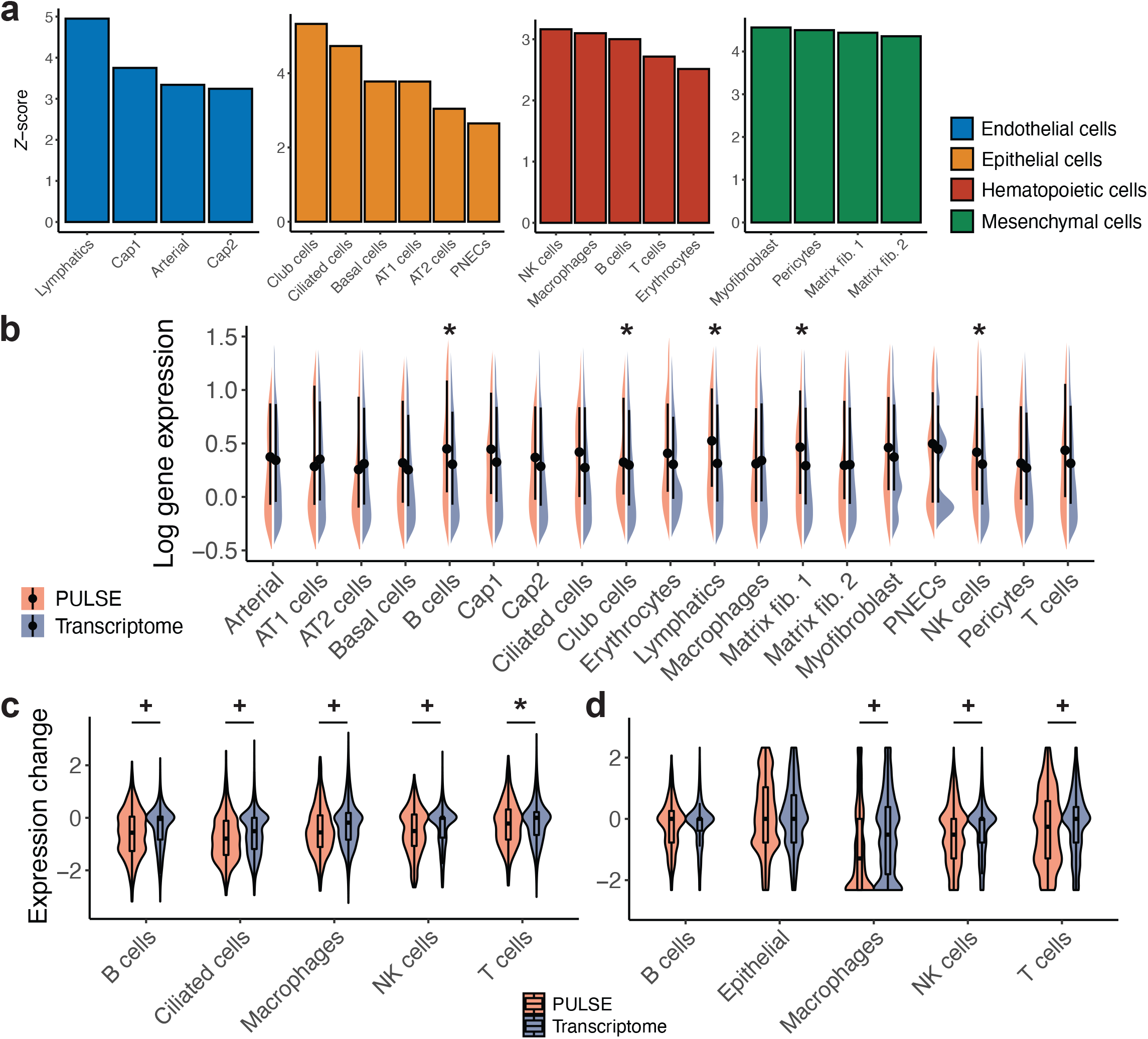
Transcriptome analysis of PULSE COVID-19 genes. **a**, Analysis of convergence between PULSE and RefMap COVID-19 genes. The *Z*-scores were calculated per cell-type by Wilcoxon rank-sum test of the difference in PULSE weights between RefMap genes and the background transcriptome. Non-zero *Z*-scores indicate biological overlap between common and rare variant architectures detected by RefMap and PULSE, respectively. **b**, Gene expression analysis of PULSE genes across different cell types in healthy lungs. The transcriptome was defined as the total set of expressed genes for each cell type (**Methods**). Violin plots show the distributions of log expression levels within each group, and point plots indicate the median and IQR. **c, d**, Comparative gene expression analysis of cell-type-specific PULSE genes in severe COVID-19 patients versus moderate patients based on scRNA-seq datasets from (**c**) Ren et al. and (**d**) Liao et al., respectively. The *Z*-score of Wilcoxon rank-sum test was used to indicate the gene expression change between severe and moderate patient groups. Violin plots show the distribution of gene expression changes within each group, and boxplots indicate the median, IQR, Q1−1.5×IQR, and Q3+1.5×IQR. *: FDR<0.1. +: FDR<0.01.

To further validate the importance of genes captured by PULSE for severe COVID-19, we identified 657 genes (referred to as PULSE COVID-19 genes) based on model weights in the top 5% from all genes (**Supplementary Table 10**). We re-examined our SKAT burden analysis results for the GEN-COVID cohort and observed that PULSE genes are significantly enriched with severe-disease-associated rare variants (median *P*<1e-5, permutations test). Similar enrichment was confirmed based on an independent rare-variant burden analysis from Regeneron^12^, where the PULSE genes are significantly enriched with Regeneron genes implicated in the analysis of severe COVID-19 versus non-hospitalized COVID-19 (*n*=68 genes; *P*=0.02, OR=2.9, Fisher’s exact test; **Methods**).

To gain further insights into the cell-type-specificity of PULSE genes, we investigated their expression levels in healthy lungs per cell type. We confirmed the function of PULSE genes across different cell types by observing their non-random overlapping with lung snATAC-seq peaks^33^ (FDR<0.1, permutation test). Furthermore, the expression levels of PULSE genes measured by scRNA-seq were examined, where we found that PULSE genes are higher expressed in B cells, club cells, lymphatics, matrix fibroblast 1, and NK cells (FDR<0.1, one-tailed Wilcoxon rank-sum test; **Fig. 5b**). This supports the functional importance of PULSE genes in lung function.

PULSE genes carry a higher mutation burden in severe COVID-19 and therefore we hypothesized that loss of function of PULSE genes leads to severe symptoms. To validate this, we analyzed the expression levels of PULSE genes based on the scRNA-seq data of COVID-19 patients^23^. Consistent with our hypothesis, we observed a down-regulation of PULSE genes in severe disease compared to moderate disease across B cells, ciliated cells, macrophages, NK cells, and T cells (FDR<0.1, one-tailed Wilcoxon rank-sum test; **Fig. 5c**). A similar analysis in another cohort^22^ led to the same conclusion for macrophages, NK cells, and T cells (FDR<0.1, one-tailed Wilcoxon rank-sum test; **Fig. 5d**). Our transcriptome study demonstrates the functional role of PULSE genes in severe disease across multiple cell types. Notably, among all the cell types we investigated, only NK cells are consistently associated with severe COVID-19 across all observations. This supports the conclusion of our common variant analysis, suggesting that NK cells are vital determinants of COVID-19 severity.

### Systems analysis implicates association of NK cell activation with COVID-19 severity

All of our analyses have suggested that NK cell dysfunction is a determinant of COVID-19 severity. To obtain a comprehensive landscape of NK cell biology underlying severe COVID-19, we examined the function of NK-cell genes identified by either RefMap or PULSE (377 genes; **Supplementary Table 11**). Indeed, genes do not function in isolation^58,59^ and therefore, rather than examining individual genes, we mapped NK-cell genes to the global protein-protein interaction (PPI) network and then inspected functional enrichment of COVID-19-associated network modules.

In particular, we extracted high-confidence (combined score >700) PPIs from STRING v11.0^60^, which include 17,161 proteins and 839,522 protein interactions. To eliminate the bias of hub genes^61^, we performed the random walk with restart algorithm over the raw PPI network to construct a smoothed network based on edges with weights in the top 5% (**Supplementary Table 12**; **Methods**). Next, this smoothed PPI network was decomposed into non-overlapping subnetworks using the Leiden algorithm^62^. This process yielded 1,681 different modules (**Supplementary Table 13**), in which genes within modules are densely connected but sparsely connected with genes in other modules.

NK-cell COVID-19 genes were mapped to individual modules, and four modules were found to be significantly enriched with NK-cell genes: M237 (*n*=471 genes; FDR<0.1, hypergeometric test; **Fig. 6a**), M1164 (*n*=396 genes; FDR<0.1, hypergeometric test; **Fig. 6b**), M1311 (*n*=14 genes; FDR<0.1, hypergeometric test), and M1540 (*n*=226 genes; FDR<0.1, hypergeometric test; **Fig. 6c**) (**Supplementary Table 13**). We excluded M1311 from our downstream analysis due to its limited size and lack of functional enrichment.

**Figure 6.**
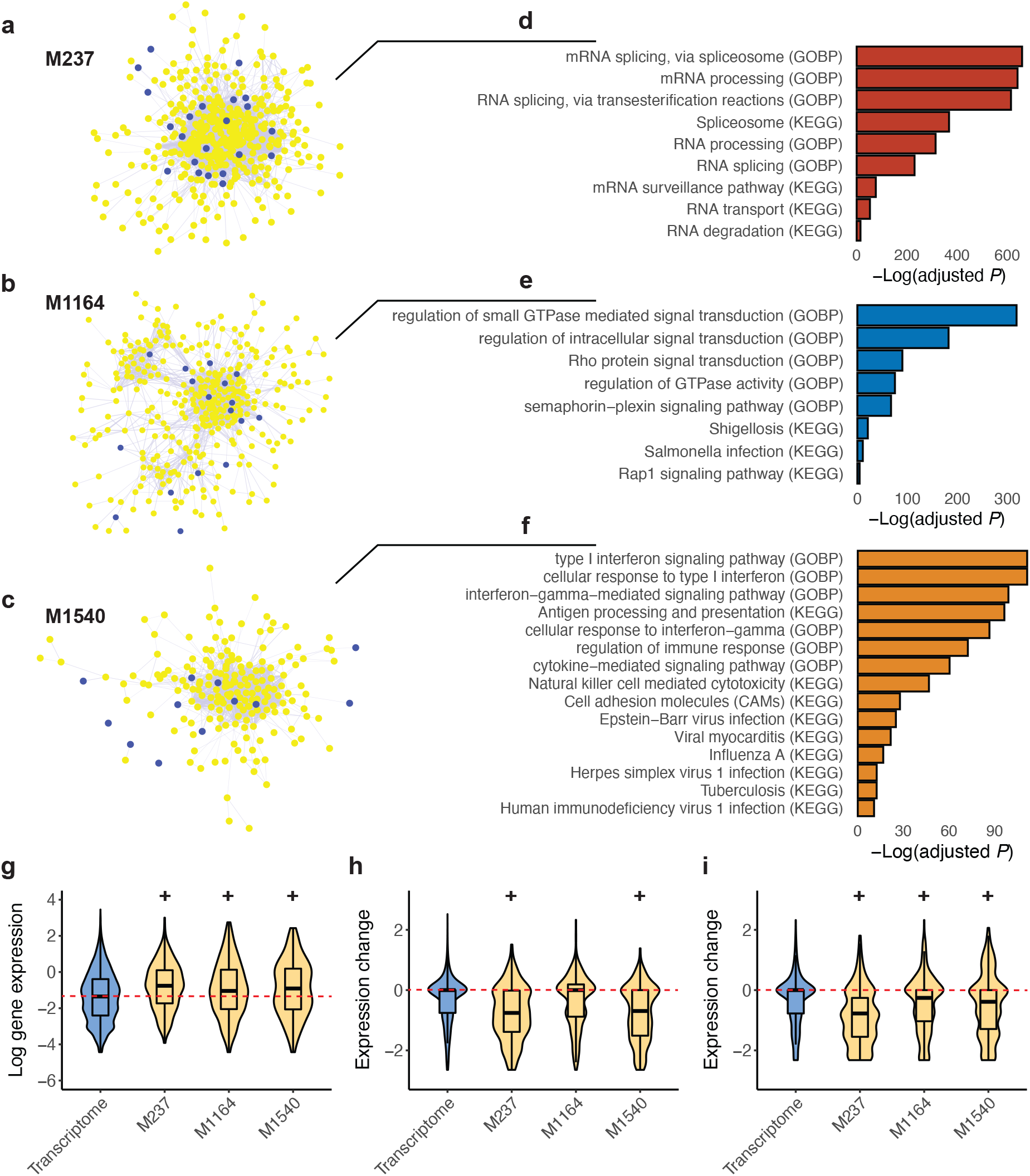
Network analysis of NK-cell genes identified in common and rare variant analyses. **a, b, c**, Three PPI network modules, including (**a**) M237, (**b**) M1164, and (**c**) M1540, are significantly enriched with NK-cell genes identified in either common or rare variant analysis. Blue nodes represent NK-cell genes and yellow nodes indicate other genes within each module. Edge thickness is proportional to STRING confidence score (>700). **d, e, f**, Gene Ontology (GO) terms that are significantly enriched in modules (**d**) M237, (**e**) M1164, and (**f**) M1540. Selected terms are shown for visualization and the complete lists can be found in **Supplementary Tables 14** and **15. g**, Gene expression analysis of module genes in NK cells. The transcriptome was defined as the total set of expressed genes in NK cells (**Methods**). Violin plots show the distributions of log expression levels within each group, and boxplots indicate the median, IQR, Q1−1.5×IQR, and Q3+1.5×IQR. The red dashed line indicates the median expression level of the transcriptome. **h, i**, Comparative gene expression analysis of module genes in severe COVID-19 patients versus moderate patients based on scRNA-seq datasets from (**h**) Ren et al. and (**i**) Liao et al., respectively. The *Z*-score of Wilcoxon rank-sum test was used to indicate the gene expression change between severe and moderate patient groups. Violin plots show the distribution of gene expression changes within each group, and boxplots indicate the median, IQR, Q1−1.5×IQR, and Q3+1.5×IQR. The red dashed line indicates the median expression change of the transcriptome. +: FDR<0.01. GOBP: gene ontology biological process.

Functionally, M237, M1164, and M1540 are all enriched with gene expression linked to NK cells (*P*<0.05, Human Gene Atlas), demonstrating their specificity in the NK cell function. Moreover, these three modules relate to different stages of NK cell activation. M237 is enriched with GO/KEGG terms including ‘mRNA processing (GO:0006397)’ and ‘Spliceosome’, which are important for the transcriptional response involved in NK cell activation (adjusted *P*<0.1; **Fig. 6d, Supplementary Tables 14** and **15**). M1164 is enriched with GO/KEGG terms linked to intracellular signalling (e.g., ‘regulation of small GTPase mediated signal transduction (GO:0051056)’), including pathways (e.g., ‘Rap1 signalling pathway’) key for NK cell activation (adjusted *P*<0.1; **Fig. 6e, Supplementary Tables 14** and **15**). M1540 is highly enriched with GO/KEGG terms linked to type I interferon signalling (e.g., ‘type I interferon signalling pathway (GO:0060337)’ and ‘Antigen processing and presentation’) (adjusted *P*<0.1; **Fig. 6f, Supplementary Tables 14** and **15**). In summary, the functional enrichment of M237, M1164, and M1540 genes includes extracellular, cytoplasmic, and nuclear processes necessary for NK cell activation; thus the genetic architecture we have discovered places NK cell activation upstream in determining severe COVID-19.

To further characterize the function of the identified NK-cell modules, we investigated the expression of module genes based on scRNA-seq data from healthy and diseased lung tissues. Genes in all three modules are relatively over-expressed in NK cells of healthy lungs^33^ than the background transcriptome (FDR<0.01, one-tailed Wilcoxon rank-sum test; **Fig. 6g**). In contrast, in lung tissues infected with SARS-CoV-2, we observed a down-regulation of M237 and M1540 genes in NK cells of severe disease^23^ (FDR<0.01, one-tailed Wilcoxon rank-sum test; **Fig. 6h**). M1164 genes are also down-regulated in NK cells from severe COVID-19 patients in another cohort^22^ (FDR<0.01, one-tailed Wilcoxon rank-sum test; **Fig. 6i**) along with M237 and M1540 genes. These results are consistent with our previous findings and functionally link the modules we have detected to NK cell biology in the context of severe COVID-19.

## DISCUSSION

The COVID-19 pandemic is a global health crisis^1^. Vaccination efforts have led to early successes^63^, but the prospect of evolving variants capable of immune-escape^64^ highlights the importance of efforts to better understand the COVID-19 pathogenesis and to develop effective treatments. Host genetic determinants of disease severity have been investigated^5–12^, but the findings and functional interpretations so far have been limited^13^. In contrast, studies of the immune response accompanying severe COVID-19^16–19^ have struggled to establish causality leading to a diverse array of candidates and little consensus. Our contribution is an integrated analysis of common and rare host genetic variation causally linked to severe COVID-19 in non-elderly adults, together with biological interpretations via single-cell omics profiling of lung tissue, and identification of >1,000 risk genes.

Our study of common and rare genetic variation associated with severe COVID-19 converges on common biology, despite non-overlapping datasets and orthogonal analytical methods. We have achieved this because we have developed effective machine learning methods which offer advantages over traditional methods: RefMap to integrate common variants with epigenetic profiles^32^, and PULSE for rare variant discovery by prediction. The evolution of clinical COVID-19 involves the interaction of multiple viral and host factors in what is likely to be a nonlinear system; our work supports this proposal and suggests that traditional methods may be inadequate given current sample sizes. This study is the first time we have presented PULSE and we have demonstrated a significant power advantage compared to standard methodology. Both methods are ready for application in other disease areas.

Our network analysis highlights NK cell activation through type I interferon signalling (**Fig. 6f**) as a key upstream determinant of COVID-19 severity. This links to previous literature describing a delayed interferon response as a precursor of later hyperinflammation associated with potentially fatal ARDS^27,65^. NK cells can also be activated via MHC signalling through NKG2 proteins. The CD94/NKG2C/HLA-E axis has been shown to be key to the NK antiviral response^66^ but so has the recognition of induced-self antigens via the NKG2D receptor^67^. Deletions of NGK2C have previously been linked to severe COVID-19^28^, whereas both our Mendelian randomization and transcriptome analyses highlight a role for NKG2D+ NK cells. We suggest that all three mechanisms for NK cell activation are critical to the host immune response to SARS-CoV-2. Indeed, a recent study has revealed that autoantibodies which impair NK cell activation are associated with severe COVID-19, and that manipulating the activation of NK cells in a mouse model resulted in a significantly higher viral burden^29^. In the cancer field, NK cell stimulation has been postulated as a therapeutic strategy^68^. We propose that this strategy could protect at-risk individuals in future waves of COVID-19.

It is important to note that our analyses also identified genetic risk of severe COVID-19 associated with non-NK cell types, including other immune cells and epithelial cells such as AT2 cells, which is consistent with the previous literature^69^. Indeed, the PULSE prediction is based on a total genetic architecture and not limited to NK cell genomics. Future work will determine how these other cell types are essential and how they interact with NK cell activation.

We present a validated prediction of COVID-19 severity derived entirely from host characteristics, including age, sex, and genetics. The average AUROC of ∼0.72 outperforms all comparable strategies^9^; and we achieve a very high sensitivity of ∼85% with a specificity >50%. Our prediction could be applied in advance of infection or even exposure, and thus has the potential to be very useful clinically. We anticipate future use and refinement of our prediction model to guide administration of post-exposure prophylaxis to at-risk individuals, in a similar manner to current standard practice for HIV^70^.

Our analyses are based on the largest available datasets to date but increasing sample size could improve the precision of our discovery and prediction. In addition, the vast majority of our data was taken from populations and at times when recently identified SARS-CoV-2 variants were not prevalent in the population (before November 2020, https://covariants.org/per-country). It is unlikely, but not impossible, that the NK cell responses we have identified as essential determinants of severe COVID-19 are not applicable to new variants.

In conclusion, we have uncovered a comprehensive genetic architecture of severe COVID-19 integrated with single-cell-resolution biological functions. Both common and rare variant analyses have highlighted NK cell activation as a potential key factor in determining disease severity. Our novel rare variant method has also achieved age-, sex-, and ancestry-independent prediction of COVID-19 severity from personal genomes.

## METHODS

### The RefMap model

Allele *Z*-scores were calculated as *Z*=*b*/*se*, where *b* and *se* are effect size and standard error, respectively, as reported by the COVID-19 GWAS^7^ (COVID-19 Host Genetics Initiative, Release 5, phenotype definition A2, EUR only) where the sample age, sex, and ancestry information were included as covariates. Given *Z*-scores and lung snATAC-seq peaks, we aim to identify functional genomic regions in which the *Z*-score distribution is significantly shifted from the null distribution. Suppose we have *K* 1Mb linkage disequilibrium (LD) blocks, where each LD block contains *J*_*k*_(*k*=*1*, …, *K*) 1kb regions and each region harbors *I*_*j,k*_(*j*=*1*, …, *J*_*k*_, *I*_*j,k*_>0) SNPs, the *Z*-scores follow a multivariate normal distribution, i.e.,

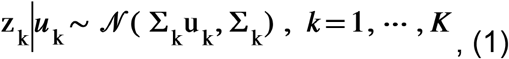

in which the *Z*-score of the *i*-th SNP in the *j*-th region of the *k*-th block is denoted as *z*_*i,j,k*_ (*i*=*1*, …, *I*_*j,k*_) and ***u***_*k*_ are the effect sizes that can be expressed as

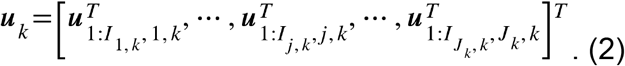

In addition, 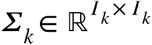 in Eq. (1) represents the in-sample LD matrix comprising of the pairwise Pearson correlation coefficients between SNPs within the *k*-th block, where *I*_*k*_ is the total number of SNPs calculated by 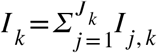. Here, since we have no access to the individual-level data, we used EUR samples from the 1000 Genomes Project (Phase 3) to estimate *Σ*_*k*_, yielding the out-sample LD matrix. A modified Cholesky algorithm^71^ was used to get a symmetric positive definite (SPD) approximation of the LD matrix.

Further, we assume *u*_*i,j,k*_ (*i*=*1*, …, *I*_*j,k*_) are independent and identically distributed (i.i.d.), following a normal distribution given by

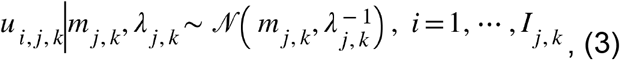

where the precision *λ*_*j,k*_ follows a Gamma distribution, i.e.,

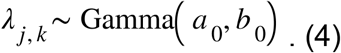

Moreover, to characterize the shift of the expectation in Eq. (3) from the null distribution, we model *m*_*j,k*_ by a three-component Gaussian mixture model given by

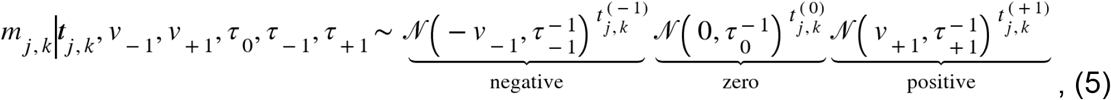

where the precisions follow

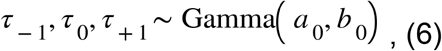

and *v*_*-1*_ and *v*_*+1*_ are non-negative variables measuring the absolute values of effect size shifts for the negative and positive components, respectively.

To impose non-negativity over *v*_*-1*_ and *v*_*+1*_, we adopt the rectification nonlinearity technique proposed previously^72^. In particular, we assume *v*_*-1*_ and *v*_*+1*_ follow

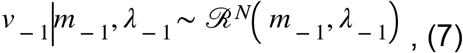

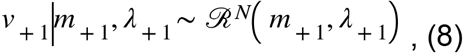

in which the rectified Gaussian distribution is defined via a dumb variable. In particular, we first define *v*_*-1*_ and *v*_*+1*_ by

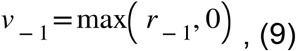

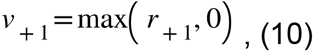

which guarantees that *v*_*-1*_ and *v*_*+1*_ are non-negative. The dump variable *r*_*-1*_ and *r*_*+1*_ follow the Gaussian distributions given by

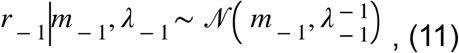

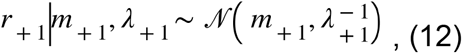

where *m*_±_ and *λ*_±_ follow the Gaussian-Gamma distributions, i.e.,

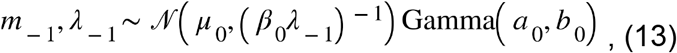

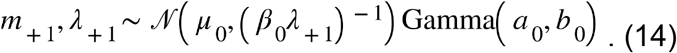

The indicator variables in Eq. (5) denote whether that region is disease-associated or not. Indeed, we define the region to be disease-associated if 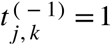 or 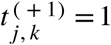, and to be non-associated otherwise. To simplify the analysis, we put a symmetry over 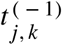 and 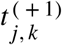, and define the distribution by

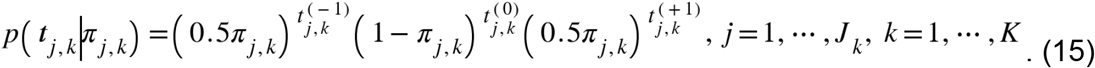

Furthermore, the probability parameter *π*_*j,k*_ in Eq. (15) is given by

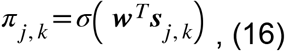

Where *σ*(.) is the sigmoid function, ***S***_*j,k*_ is the vector of epigenetic features for the *j*-th region in the *k*-th LD block, and the weight vector ***w*** follows a multivariate normal distribution, i.e.,

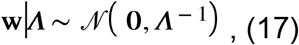

and *⋀* follows

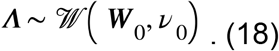

In this study, the epigenetic feature ***s***_*j,k*_ was calculated as the overlapping ratios of that region with the snATAC-seq peaks detected in any of the cell types in healthy human lungs.

Based on the model defined in Eqs. (1) to (18), we are interested in calculating the posterior probability *p*(***T*** | ***Z, S***), where the mean-field variational inference (MFVI)^73^ was adopted to solve the intractability. More technical details, including a coordinate ascent-based inference algorithm, can be found in our previous work^32^.

In this study, we ran the MFVI algorithm per chromosome to accelerate the computation. The *Q*^*+*^- and *Q*^*-*^-scores were defined as *q*(*t*^(+ 1)^ = 1) and *q*(*t*^(− 1)^ = 1), respectively, and we also defined the *Q*-score as *Q*=*Q*^*+*^+*Q*^*-*^. RefMap regions were identified by *Q*^*+*^- or *Q*^*-*^-score >0.95.

### Mapping cell-type-specific genes from RefMap regions

For each cell type within lung tissue, we defined cell-type-specific RefMap regions as the overlap between RefMap regions and the total set of snATAC-seq peaks detected in that cell type (**Supplementary Table 1**). Cell-type-specific RefMap genes were then identified if the extended gene body (i.e., the region up to 10kb either side of the annotated gene body) overlapped with any of the cell-type-specific regions. To get the final gene lists, RefMap genes were further filtered based on their expression levels. In particular, with the lung snRNA-seq data^33^, we defined expressed genes in each cell type as those with Seurat^74^ log-normalized value>0.6931. In addition, we note that there are non-adult samples (∼30 weeks gestation and ∼3 years) sequenced in the single cell profiling data^33^. To remove the bias towards lung development, we first calculated the fold change of gene expression levels between the adult sample (∼30 years) and non-adult ones, and defined non-developmental genes (nDG) as those with FC>1.5. Only RefMap genes that were identified as expressed and non-developmental in each cell type were kept for downstream analysis (**Supplementary Table 1**).

### Validation of RefMap COVID-19 regions in the 23andMe dataset

We calculated the overlap of total RefMap regions and of cell-type-specific RefMap regions with genomic regions shown to contain COVID-19-associated SNPs (*P*<1e-04) based on the GWAS of an independent cohort recruited by 23andMe^5^. To determine whether the observed overlap is statistically significant, we examined the average overlap with ten sets of control regions of equivalent length to RefMap regions. Control regions were +/-1Mb-5Mb distant from the RefMap regions^75^.

### Heritability analysis

We used LD score regression (LDSC)^34^ to calculate overall heritability for severe COVID-19 (A1), hospitalized COVID-19 (B2), and COVID-19 overall (C2), respectively. Heritability partitioning within genes identified by traditional methods and within cell-type-specific RefMap genes was performed as previously described^76^. Briefly, for all gene lists, we examined the proportion of total SNP-based heritability carried by SNPs +/-100kb from the transcription start site (TSS) of each gene in the list. Enrichment was calculated by comparing the ratio of partitioned heritability to the quantity of genetic materials.

### Mendelian randomization

In total, 46 GWAS measures of NK cell subtypes were identified from the IEU Open GWAS Project, including “prot-a-180”, “met-b-124”, “met-b-245”, “met-b-242”, “prot-c-5244_12_3”, “met-b-237”, “met-b-258”, “prot-a-1669”, “prot-c-2917_3_2”, “met-b-246”, “prot-a-1671”, “met-b-249”, “met-b-140”, “met-b-240”, “prot-a-3159”, “prot-c-5104_57_3”, “prot-c-3056_11_1”, “prot-a-13”, “prot-a-3160”, “met-b-123”, “met-b-250”, “met-b-239”, “met-b-120”, “met-b-154”, “prot-a-3162”, “met-b-247”, “met-b-251”, “met-b-238”, “met-b-243”, “prot-a-2487”, “met-b-244”, “prot-c-2734_49_4”, “met-b-153”, “prot-a-3161”, “prot-c-3003_29_2”, “met-b-248”, “prot-a-1674”, “prot-a-1675”, “met-b-152”, “met-b-122”, “met-b-121”, “prot-a-1670”, “prot-c-5424_55_3”, “met-b-252”, “prot-a-3233” and “met-b-241”^37,77,78^. Exposure SNPs or instrumental variables (IVs) are chosen based on an arbitrary *P*-value cutoff^79,80^. A cutoff that is too low will lose informative instruments, but a cutoff that is too high could introduce non-informative instruments. We chose to set the cutoff at 5e-06 in line with our previous work^81^. We employed a series of sensitivity analyses to ensure that our analysis was not confounded by invalid IVs. Identified SNPs were clumped for independence using PLINK clumping in the TwoSampleMR tool^82^. A stringent cutoff of *R*^2^≤0.001 and a window of 10,000kb were used for clumping within a European reference panel. Where SNPs were in LD, those with the lowest *P*-value were retained. SNPs that were not present in the reference panel were excluded. Where an exposure SNP was unavailable in the outcome dataset, a proxy with a high degree of LD (*R*^2^≥0.9) was identified in LDlink within a European reference population^83^. Where a proxy was identified to be present in both datasets, the target SNP was replaced with the proxy in both exposure and outcome datasets in order to avoid phasing issues^84^. Where a SNP was not present in both datasets and no SNP was available in sufficient LD, the SNP was excluded from the analysis. The effects of SNPs on outcomes and exposures were harmonized in order to ensure that the beta values were signed with respect to the same alleles. For palindromic alleles, those with minor allele frequency (MAF) > 0.42 were omitted from the analysis in order to reduce the risk of errors due to strand issues^84^.

The MR measure with the greatest power is the inverse-variance weighted (IVW) method, but this is contingent upon the exposure IV assumptions being satisfied ^85^. With the inclusion of a large number of SNPs within the exposure IV, it is possible that not all variants included are valid instruments and therefore, in the event of a significant result, it is necessary to include a range of robust methods which provide valid results under various violations of MR principles at the expense of power^86^. Robust methods applied in this study include MR-Egger, MR-PRESSO, weighted median, weighted mode, and MR-Lasso.

With respect to the IVW analysis, a fixed-effects (FE) model is indicated in the case of homogeneous data, whilst a multiplicative random effects (MRE) model is more suitable for heterogeneous data. Burgess et al. recommended that an MRE model be implemented when using GWAS summary data to account for heterogeneity in variant-specific causal estimates^86^. In the interest of transparency, we calculated both results but present the MRE in the text.

MR analyses should include evaluation of exposure IV strength. In order to achieve this, we provided the *F*-statistic, MR-Egger intercept, MR-PRESSO global test, Cochran’s *Q* test, and *I*^2^ for our data. The *F*-statistic is a measure of instrument strength with >10 indicating a sufficiently strong instrument^87^. We provided *F*-statistics for individual exposure SNPs and the instrument as a whole. Cochran’s *Q* test is an indicator of heterogeneity in the exposure dataset and serves as a useful indicator that horizontal pleiotropy is present as well as directing decisions to implement FE or MRE IVW approaches^88^. The MR-Egger intercept test determines whether there is directional horizontal pleiotropy. The MR-PRESSO global test determines if there are statistically significant outliers within the exposure-outcome analysis^89^. *I*^2^ was calculated as a measure of heterogeneity between variant specific causal estimates, with a low *I*^2^ indicating that Egger is more likely to be biased towards the null^90^. Finally, we performed a leave-one-out analysis using the method of best fit for each exposure SNP within the IV in order to determine if any single variants were exerting a disproportionate effect upon the results of our analysis^86^.

### MAGMA analysis of COVID-19 GWAS data

MAGMA (v1.08)^91^ was applied using default settings. Input consisted of summary statistics for all SNPs genome-wide as measured in the COVID-19 GWAS^7^. We estimated LD structure using EUR samples from the 1000 Genomes Project (Phase 3). The top 50 MAGMA genes^35^ were used for downstream analysis.

### DNA sequencing in rare variant analysis

#### GEN-COVID cohort

The cohort was recruited by the GEN-COVID consortium (https://sites.google.com/dbm.unisi.it/gen-covid) as described previously^52^. Briefly, adult patients (>18 years) were recruited from 35 Italian hospitals starting on March 16, 2020. Infection status was confirmed by SARS-CoV-2 viral RNA polymerase-chain-reaction (PCR) test collected at least from nasopharyngeal swabs. Demographics and clinical severity were assessed via an extensive questionnaire.

Sequencing and variant calling were performed as described previously^52^. Briefly, sample preparation was performed following the Nextera Flex for Enrichment manufacturer protocol. Whole-exome sequencing was performed with >97% coverage at 20X using the Illumina NovaSeq 6000 System (Illumina, San Diego, CA, USA). Reads were aligned to human reference genome build GRCh38 using BWA^92^. Variants were called according to the GATK4 best practice guidelines^93^. Duplicates were removed by *MarkDuplicates*, and base qualities were recalibrated using *BaseRecalibration* and *ApplyBQSR. HaplotypeCaller* was used to calculate Genomic VCF files for each sample, which were then used for multi-sample calling by *GenomicDBImport* and *GenotypeGVCF*. In order to improve the specificity-sensitivity balance, variant quality scores were calculated by *VariantRecalibrator* and *ApplyVQSR*. Variants with sequencing depth <20X were excluded.

#### VA cohort

Whole-genome sequence data on the VA COVID-19 cohort was derived from the VA Million Veteran Program (MVP). The VA MVP is an ongoing national voluntary research program that aims to better understand how genetic, lifestyle, and environmental factors influence veteran health^94^. Briefly, individuals aged 18 to over 100 years old have been recruited from over 60 VA Medical Centers nationwide since 2011 with current enrollment at >800,000. Informed consent is obtained from all participants to provide blood for genomic analysis and access to their full electronic health record (EHR) data within the VA prior to and after enrollment. The study received ethical and study protocol approval from the VA Central Institutional Review Board in accordance with the principles outlined in the Declaration of Helsinki. COVID-19 cases were identified using an algorithm developed by the VA COVID National Surveillance Tool based on reverse transcription polymerase chain reaction laboratory test results conducted at VA clinics, supplemented with natural language processing on clinical documents for SARS-CoV-2 tests conducted outside of the VA^95^.

DNA isolated from peripheral blood samples was used for whole-genome sequencing. Libraries were prepared using KAPA hyper prep kits, PCR-free according to manufacturers’ recommendations. Sequencing was performed using Illumina NovaSeq 6000 System (Illumina, San Diego, CA, USA) with paired-end 2×150bp read lengths, and Illumina’s proprietary reversible terminator-based method. The specimens were sequenced to a minimum depth of 25X per specimen and an average coverage of 30X per plate.

WGS data processing in the MVP was performed via the functional equivalence GATK variant calling pipeline^96^, which was developed by the Broad Institute and plugged into our data and task management system Trellis. The human reference genome build was GRCh38. We used BWA-MEM (v0.7.15) to align reads, Picard 2.15.0 to mark PCR duplicates, and GATK 4.1.0.0 for BQSR and variant calling via the *haplotypeCaller* function. We also used FASTQC (v0.11.4), SAMTools *flagstat* (v0.1.19), and RTG Tools *vcfstats* (v3.7.1) to assess the qualities of the FASTQ, BAM, and gVCF files, respectively. In addition, we used *verifybamID* in GATK 4.1.0.0 to estimate DNA contamination rates for individual genomes and removed samples with 5% or more contaminated reads.

### Data quality control

#### GEN-COVID cohort

To guarantee high quality of the sequencing data, we performed numerous quality control procedures. On the sample level, we (1) computed inbreeding coefficients (Fhat1, Fhat2, and Fhat3 in GCTA^97^) and removed genomes that resided more than 3 standard deviation from the mean; (2) computed identity-by-descent (IBD) and only kept one genome from pairs with proportion IBD>0.2; (3) computed missing calls for each genome and removed those with missing rate larger than 10%; (4) computed singleton calls, SNV count, indel count, Ti/Tv ratio, and heterozygous calls for each genome and removed genomes that resided more than 3 standard deviation from the mean.

On the variant level, we (1) removed multiallelic sites; (2) kept variants in autosomes; (3) removed variants on blacklisted regions, compiled by the ENCODE Project Consortium (Phase 4); (4) removed variants identified other than ‘‘PASS,’’ such as ‘‘low quality,’’ ‘‘tranche99.0-99.5,’’ by VQSR in GATK; (5) removed variants with missing rate larger than 10%. The samples which passed QCs were provided in **Supplementary Table 8**.

#### VA cohort

For deriving high-quality variants for downstream analysis, we removed samples with kinship >0.03, sample call rate <0.97, or mean sample coverage <=18X. Genomic positions resided in low complexity regions or ENCODE blacklisted regions were first removed. Next, we filtered out genotypes in individual samples that were detected with too low or too high of read coverages (DP<5 or >1500). We required all calls to have genotype quality (GQ) >=20, and for non-reference calls, sufficient portion (>0.9) of reads was required to cover the alternate alleles. In addition, we removed genomic positions with cohort-wise call rate <0.95 and computed Hardy-Weinberg equilibrium (HWE), which was required to be <1e-05 for common variants and <1e-06 for rare variants. With all these filtering completed, we assessed the sample-level genomic parameters, such as Ti/Tv ratios, het/hom ratios, and number of singletons/SNVs/INDELs, and removed any sample that fell into the tail regions of the distribution (>=3 standard deviation). The samples which passed QCs were provided in **Supplementary Table 9**.

### Ancestry analysis

We performed population admixture analysis using ADMIXURE^98^ (v1.3.0) referencing five super populations, including AFR (African), AMR (Ad Mixed American), EAS (East Asian), EUR (European), and SAS (South Asian), in the 1000 Genomes Project (Phase 3) and inferred the ancestry for each genome. For the GEN-COVID cohort, samples with >90% EUR ancestry fraction were kept in the discovery cohort. For the VA COVID-19 cohort, we relaxed the ancestry fraction cutoff to 70% for including more samples in testing. Inferred sample ancestry can be found in **Supplementary Tables 8** and **9**.

### Variant- and gene-level annotations

Genome annotation was performed by Annovar^53^ integrating multiple databases. Variant frequency was estimated using the 1000 Genomes Project (Phase 3). Nonsynonymous (missense and nonsense) variants were annotated using dbNSFP^99^ (v3.5). The mutation effect of splicing-site variants was predicted by dbscSNV^100^ (v1.1) and regSNP-intron^101^.

### Rare-variant burden testing

Rare-variant burden testing was performed to determine whether any genes were differentially enriched with rare variants between severe COVID-19 patients and non-severe COVID-19-positive controls. We utilized whole-exome sequencing data from the GEN-COVID cohort^51^, including 122 individuals aged ≤60 years who suffered severe COVID-19 requiring respiratory support, and 465 individuals aged ≥20 years who suffered non-severe COVID-19 not requiring hospitalisation. Variants were included if they altered an amino acid, were rare (MAF<1%) and absent from the EUR cohort of the 1000 Genomes Project (Phase 3). Burden was calculated using SKAT^50^ adjusted for sample imbalance using a saddlepoint approximation^102^. Sex and the first ten principal components were included as covariates. Genetic burden was compared with the complete set of coding genes; genes caring <10 variants were removed because of insufficient data. After filtering a total set of 4,280 genes were tested for severe-COVID-19-associated rare genetic variation of which 625 were also RefMap COVID-19 genes. A QQ-plot confirmed that there was no significant genomic inflation (λGC=1.1; **Supplementary Fig. 4**).

Regeneron’s burden testing results were obtained from the Regeneron results browser (https://rgc-covid19.regeneron.com/results), where only semi-significant genes (*P*<1e-03, REGENIE^103^) were available. Data consists of exome-wide association studies of various COVID-19 outcomes across 662,403 individuals (11,356 with COVID-19) aggregated from four studies: UK Biobank (UKB; *n*=455,838), AncestryDNA COVID-19 Research Study (*n*=83,930), Geisinger Health System (GHS; *n*=113,731), and Penn Medicine BioBank (PMBB; *n*=8,904). For the Regeneron study of severe COVID-19 versus non-hospitalized COVID-19, we obtained a list of 68 genes harboring disease-associated missense mutations at a significance cutoff of *P*<1e-03 in EUR samples. Overlap between PULSE genes and Regeneron gene lists was tested by Fisher’s exact test, assuming a background of 19,396 coding genes in the genome which is the total number profiled by Regeneron^12^.

### The PULSE model

#### Feature engineering

Given the variant annotations from ANNOVAR, we calculated gene-level mutation profiles for each individual. Here we only focused on rare nonsynonymous and splicing-site SNVs as well as frameshift and splicing-site indels. Rare variants were defined as those not present within 1000 Genomes Project (Phase 3) samples. For nonsynonymous and splicing-site SNVs, we calculated the accumulative mutation burdens for each gene based on individual annotations (32 in total; **Supplementary Table 7**). For indels, the number of variants was counted for frameshift and splicing-site, respectively (**Supplementary Table 7**). Consequently, the mutation profile consists of 34 features per gene per individual.

#### Mapping phenotype from genotype

Given the mutation profiles 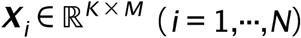 for the *i*-th sample and the corresponding disease status *y*_*i*_∈{0,1}(*y*_*i*_=1 indicates a case, and otherwise), PULSE models the conditional, which is the probability of disease status for the *i*-th sample characterized by the genome. Here *K, M*, and *N* are the numbers of annotation features, genes, and samples, respectively. Note that we have *K*=34 in this study. In particular, we aggregate the mutation profiles across the genome using a bilinear transformation and define the conditional as

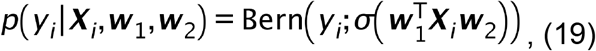

where σ(.)denotes the sigmoid function, ***w***_1_ are random variables weighing the importance of each annotation feature, and ***w***_2_ effect sizes for individual genes. We model ***w***_1_ by a multivariate Gaussian given by

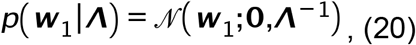

in which the precision matrix is characterized by a Wishart distribution, i.e.,

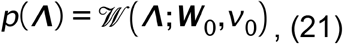

and the hyperparameters are set to ***W***_1_ = ***I***_*K*_ and v_0_ = *K* to introduce non-informative prior.

To prevent overfitting, we introduce a spike-and-slab prior over, ***w***_1_ i.e.,

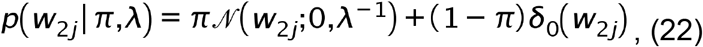

where *π* is the probability of being non-zero and *δ*_0_ (.) is the Dirac function forcing *w*_*2j*_ to be zero. Two additional conjugate priors are further used over distribution parameters in (22), i.e.,

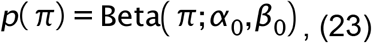

and

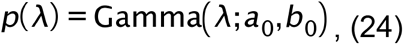

in which we set α_0_=β_0_=0.5 (i.e., the Jeffrey prior) and a_0_=b_0_=10^−6^ to keep it non-informative. In this study, to prevent false positives, accelerate computation, and eliminate the sex bias in the genetic modelling, we only considered autosomal genes that are expressed in human lungs (TPM>1 in lung RNA-seq from GTEx^104^), resulting in *M*=13,129. The diagram of the model structure is shown in Panel 3 of **Fig. 4b**.

#### Model inference

The exact inference in PULSE is intractable. Here we adopt the mean-field variational inference (MFVI), an approximate but efficient way to perform inference in Bayesian models^73^. Since the model posterior is difficult to calculate, MFVI aims to search for an optimal distribution closest to the model posterior from a family of regularized proposal distributions factorized with each other. Indeed, the solution of MFVI is given by minimizing the Kullback-Leibler (KL) divergence, i.e.,

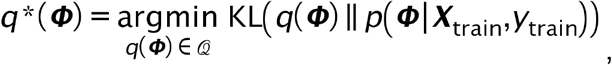

where *Φ* represents the set of hidden variables in the model, and 𝒬 is the family of factorized proposal distributions. It can be shown that minimizing the KL divergence is mathematically equivalent to maximizing the evidence lower bound (ELBO)^73^, which is solvable in optimization. Further, in order to make the MFVI for PULSE tractable and efficient, several techniques were adopted.

##### i) Local variational method

The sigmoid function in Eq. (19) makes MFVI intractable. However, instead of dealing with the sigmoid directly, we can approximately calculate the posteriors of ***w***_1_ and ***w***_2_ and with respect to its lower bound, which yields Gaussian or Gaussian-like distributions. Meanwhile, to make the approximation close to the true MFVI solution, we need to maximize the log-likelihood of observations that take the sigmoid lower bound into account with respect to local variational parameters introduced. This local variational method introduces a new objective function, which is consistent with the original MFVI. More technical details can be found in Supplementary Notes.

##### ii) Reparameterization

The spike-and-slab prior over ***w***_1_ (Eq. (22)) also makes MFVI intractable. To solve this problem, we adopted the reparameterization trick introduced in ^105^. In particular, ***w***_2_ can be reparameterized by two other variables and ***s*** and 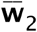, whose joint distribution is given by

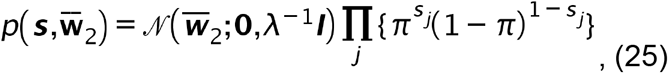

and the new variable 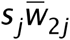 follows the same distribution as in Eq. (22). Therefore, we can do MFVI over ***s*** and 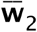 instead of ***w***_2_. However, this still introduces another problem and makes the VI highly inefficient, where the approximate posteriors from reparameterization (unimodal) could badly deviate from the original posteriors (exponentially multimodal). To alleviate this issue, a partial factorization was taken by following^105^, i.e., we assume

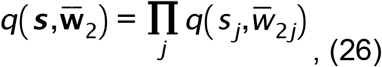

in proposal distributions, and performed MFVI over ***s***_*j*_ and 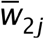 jointly. More technical details can be found in Supplementary Notes.

##### iii) Stochastic variational inference

Conventional MFVI based on coordinate ascent (i.e., CAVI) updates variational parameters in batches. However, it is difficult to deploy such a batch algorithm in big data scenarios, where the sample size or feature dimension is large. Here, stochastic variational inference (SVI)^106^ was used to scale up our model for the large amount of genome data. In fact, borrowing the idea from stochastic optimization, we can update parameters per epoch by using only one or a mini-batch of samples instead of the whole dataset. Specifically, with SVI we first calculated the natural gradient of ELBO with respect to the variational parameter whose update rule contains sample points. Thanks to the conditional conjugacy predefined in our model, the natural gradient enjoys a simple form (see Supplementary Notes for details). Then based on stochastic optimization, we sampled a minibatch and rescaled the term involving sample points, resulting in a noisy but cheaply computed and unbiased natural gradient. At last, the variational parameter was updated from this gradient according to the gradient-based optimization algorithm^107^. This SVI update can be easily embedded into CAVI without many changes. In implementation, we followed ^106^ and set the learning rate as

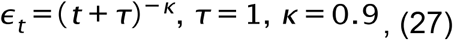

where *t* is the iteration index, Tis the delay, and k is the forgetting rate.

We integrated all above techniques into our VI algorithm. Details on the update rules for both local and global variational parameters and the VI algorithm are provided in Supplementary Notes.

#### MAP prediction

The exact Bayesian prediction for test samples needs to integrate out all hidden variables, which is computationally intense and usually not necessary. Here, we adopted maximum a posteriori (MAP) and predicted new coming sample by

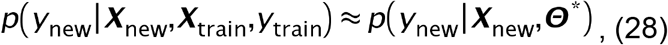

where the optimal hidden variables are given by

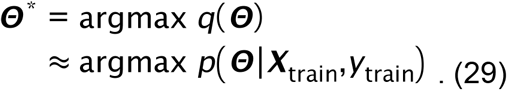

Similarly, the importance weights for individual genes (referred to as PULSE gene weights) were also estimated by the MAP of *q*(*w*_*2j*_).

### Network analysis

We first downloaded the human PPIs from STRING v11, including 19,567 proteins and 11,759,455 protein interactions. To eliminate the bias caused by hub proteins, we first carried out the random walk with restart algorithm^108^ over the PPI network, wherein the restart probability was set to 0.5, resulting in a smoothed network after retaining the top 5% predicted edges. To decompose the network into different subnetworks/modules, we performed the Leiden algorithm^62^, a community detection algorithm that searches for densely connected modules by optimizing the modularity. After the algorithm converged, we obtained 1,681 modules with an average size of 9.98 nodes (SD=53.35; **Supplementary Table 13**).

### Transcriptome analysis

Four single-cell RNA-seq datasets were used in the transcriptome analyses, including human healthy lungs^33^,39 and COVID-19 patients^22,23^. Data after QC was acquired for each study. Only samples from the respiratory system were considered in the analyses. For the healthy lung data, a cutoff of 0.6931 was used to define expressed genes in the transcriptome throughout the study if not specified. For the disease samples, we removed the overlap of severe patients between the two cohorts^22,23^. In the comparative expression analysis of severe versus moderate patients, to stabilize the analysis we estimated the change of gene expression levels using the *Z*-score estimated from Wilcoxon rank-sum test, wherein a positive *Z*-score indicates a higher expression level in severe patients and a negative value suggests the lower expression. The Benjamini-Hochberg (BH) procedure was used for multiple testing correction throughout the study.

## Data Availability

The GEN-COVID WES and clinical data are available by consultation (A.R.). The VA WGS and clinical data are available upon request from the corresponding authors (P.S.T. and M.P.S.); these data are not publicly available due to US Government and Department of Veteran's Affairs restrictions relating to participant privacy and consent. All other data used in this study are available from the original studies.

## ACKNOWLEDGEMENTS

We acknowledge the Stanford Genetics Bioinformatics Service Center (GBSC) for providing computational infrastructure for this study. This study was also supported by the National Institutes of Health (1S10OD023452-01 to GBSC; CEGS 5P50HG00773504, 1P50HL083800, 1R01HL101388, 1R01-HL122939, S10OD025212, P30DK116074, and UM1HG009442 to M.P.S.), the Wellcome Trust (216596/Z/19/Z to J.C.K.), and Million Veteran Program, Office of Research and Development, Veterans Health Administration (MVP001). This publication does not represent the views of the Department of Veteran Affairs or the United States Government. Figures 1a and 4b were created with BioRender.com.

This study is part of the GEN-COVID Multicenter Study (https://sites.google.com/dbm.unisi.it/gen-covid), the Italian multicenter study aimed at identifying the COVID-19 host genetic bases. Specimens were provided by the COVID-19 Biobank of Siena, which is part of the Genetic Biobank of Siena, member of BBMRI-IT, of Telethon Network of Genetic Biobanks (project no. GTB18001), of EuroBioBank, and of RDConnect. We thank the CINECA consortium for providing computational resources and the Network for Italian Genomes (NIG) (http://www.nig.cineca.it) for its support. We thank private donors for the support provided to A.R. (Department of Medical Biotechnologies, University of Siena) for the COVID-19 host genetics research project (D.L n.18 of March 17, 2020). We also thank the COVID-19 Host Genetics Initiative (https://www.covid19hg.org/), MIUR project “Dipartimenti di Eccellenza 2018-2020” to the Department of Medical Biotechnologies University of Siena, Italy, and “Bando Ricerca COVID-19 Toscana” project to Azienda Ospedaliero Universitaria Senese. We also thank Intesa San Paolo for the 2020 charity fund dedicated to the project “N. B/2020/0119 Identificazione delle basi genetiche determinanti la variabilità clinica della risposta a COVID-19 nella popolazione italiana” and “Bando FISR 2020” in COVID-19 from Italian Ministry of University e Research.

## AUTHOR CONTRIBUTIONS

S.Z., J.C.K. and M.P.S. conceived and designed the study. S.Z. contributed to the design, implementation, training and testing of RefMap and PULSE. S.Z., J.C.K., A.K.W., C.H., T.H.J., S.F., E.F., F.F., A.R., C.P., J.S., P.B.R., P.S.T. and M.P.S. were responsible for data acquisition. S.Z., J.C.K., C.H., T.H.J., C.W., J.L. and C.P. were responsible for data analysis. S.Z., J.C.K., A.K.W., C.H., T.H.J., C.W., J.L., S.F., E.F., F.F., A.R., C.P., P.G., X.S., I.S.T., K.P.K., M.M.D., P.S.T. and M.P.S. were responsible for the interpretation of the findings. S.Z., J.C.K., P.S.T. and M.P.S. drafted the manuscript with assistance from all authors. All authors meet the four ICMJE authorship criteria, and were responsible for revising the manuscript, approving the final version for publication, and for accuracy and integrity of the work.

## SUPPLEMENTARY INFORMATION

### SUPPLEMENTARY FIGURES

**Supplementary Figure 1.**
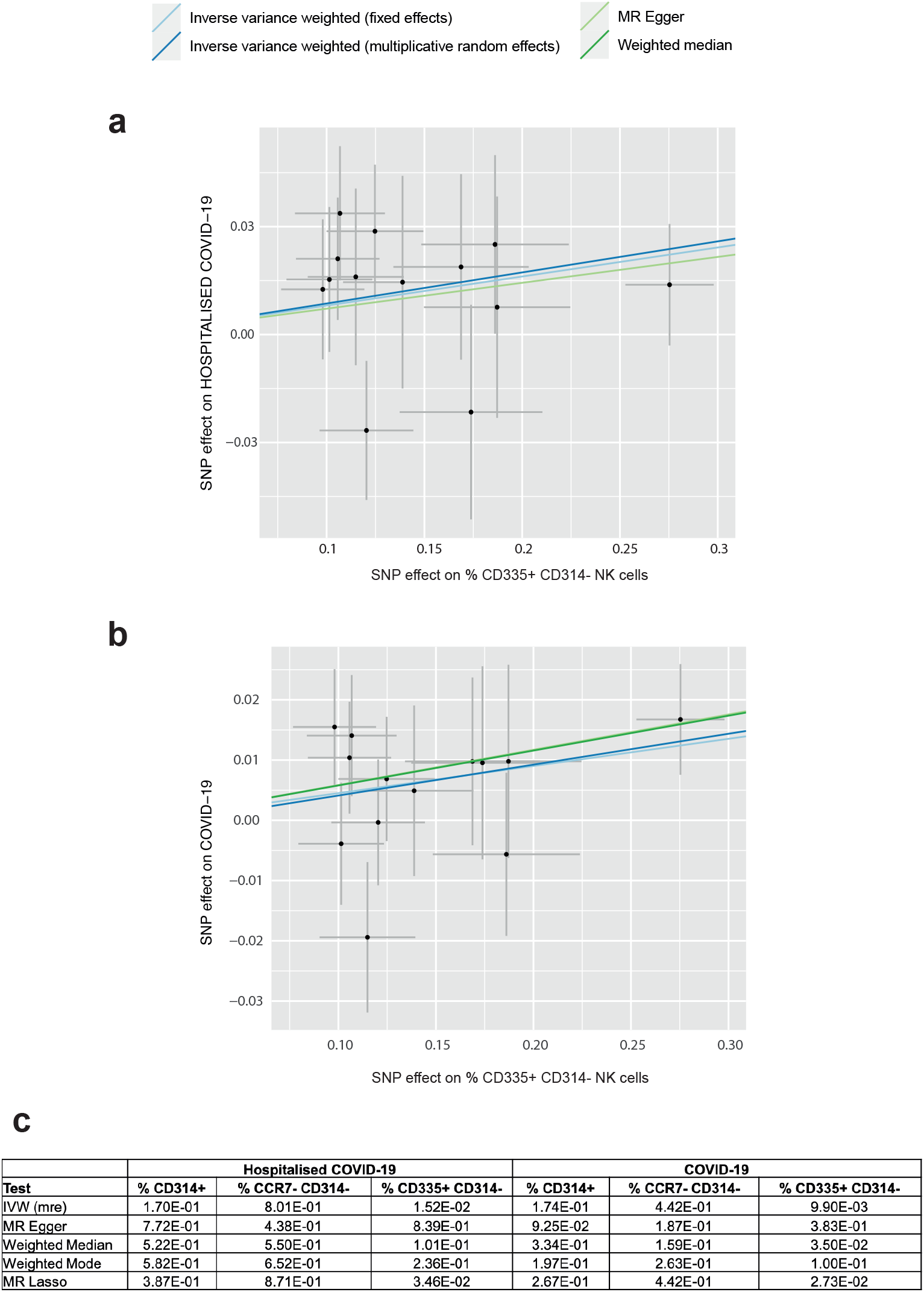
Mendelian randomization for COVID-19 GWAS with phenotypes B2 and C2

**Supplementary Figure 2.**
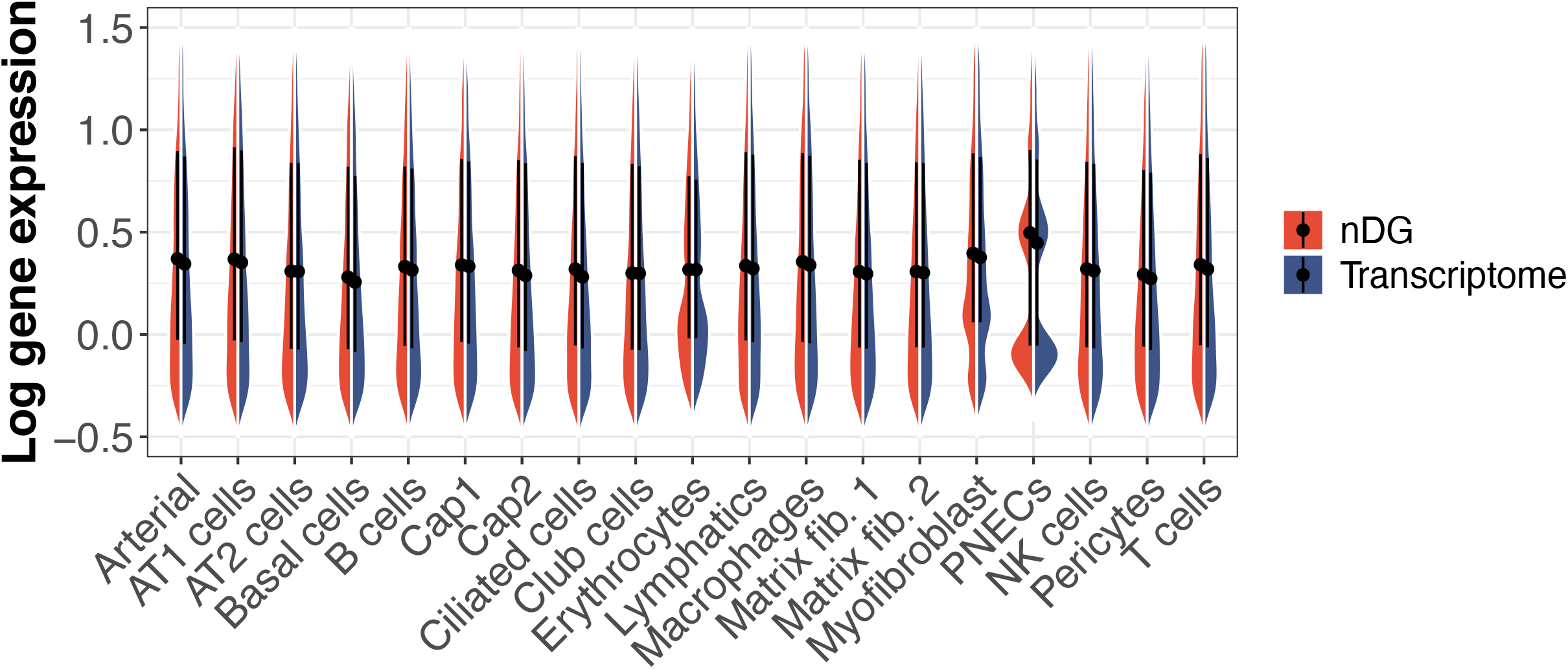
Expression levels of non-developmental genes in healthy lungs. nDG is short for non-developmental gene

**Supplementary Figure 3.**
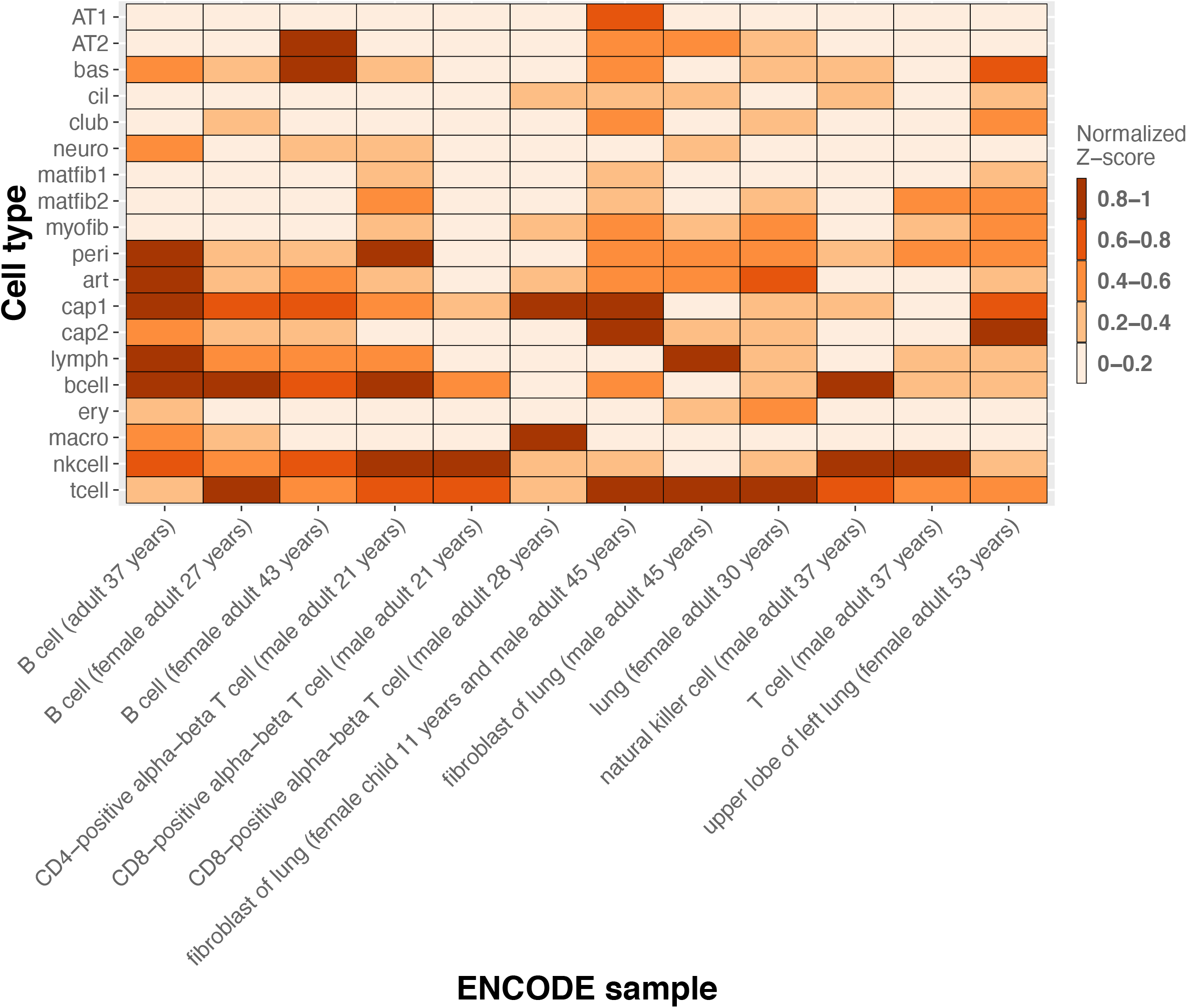
Overlap between lung snATAC-seq peaks and ENCODE ChIP-seq peaks

**Supplementary Figure 4.**
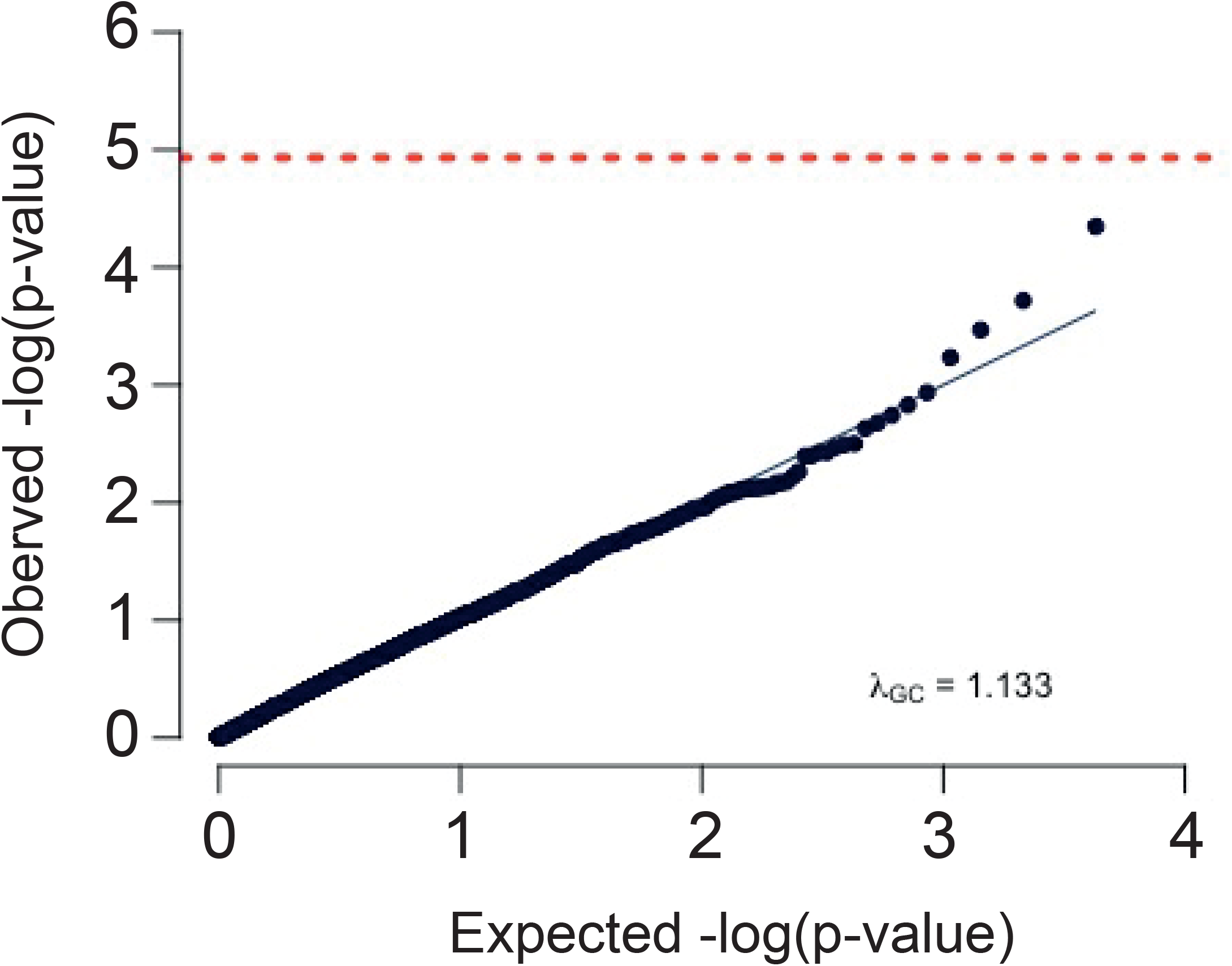
Q-Q plot of *P*-value distribution for SKAT analysis on the GEN-COVID cohort

**Supplementary Figure 5.**
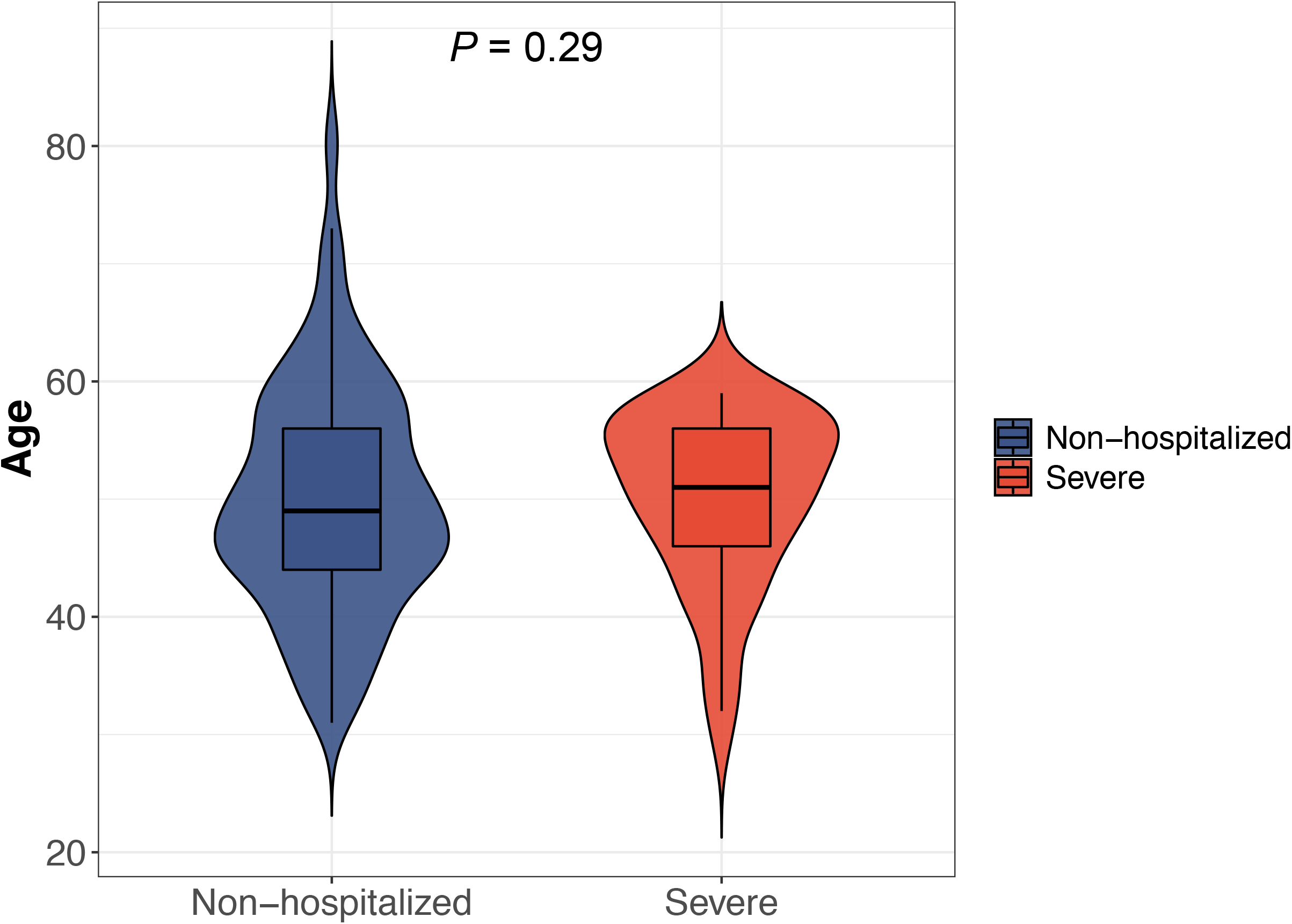
Age distribution of the GEN-COVID cohort after sample filtering

**Supplementary Figure 6.**
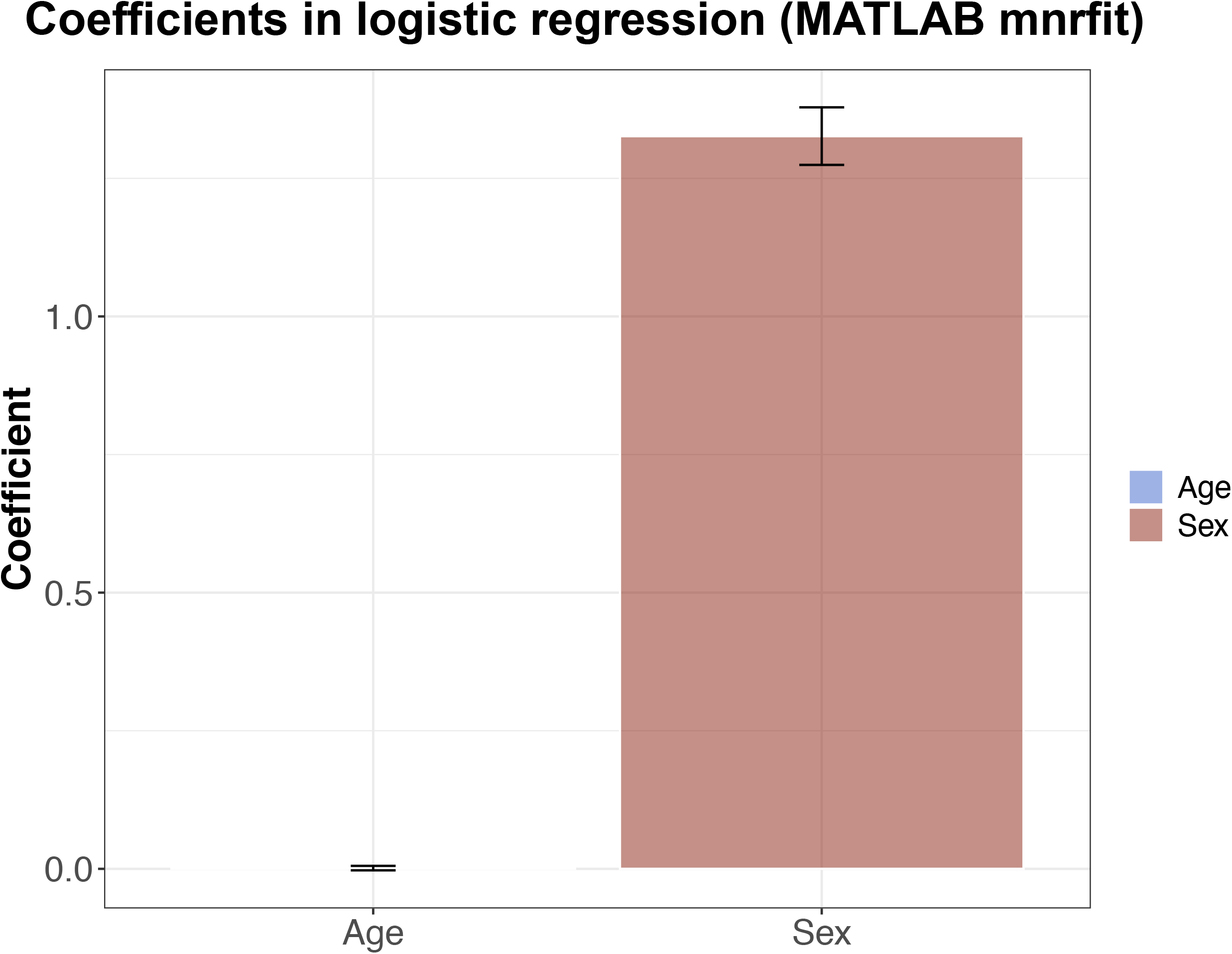
Coefficients of the age+sex logistic regression models in 5-fold cross-validation

**Supplementary Figure 7.**
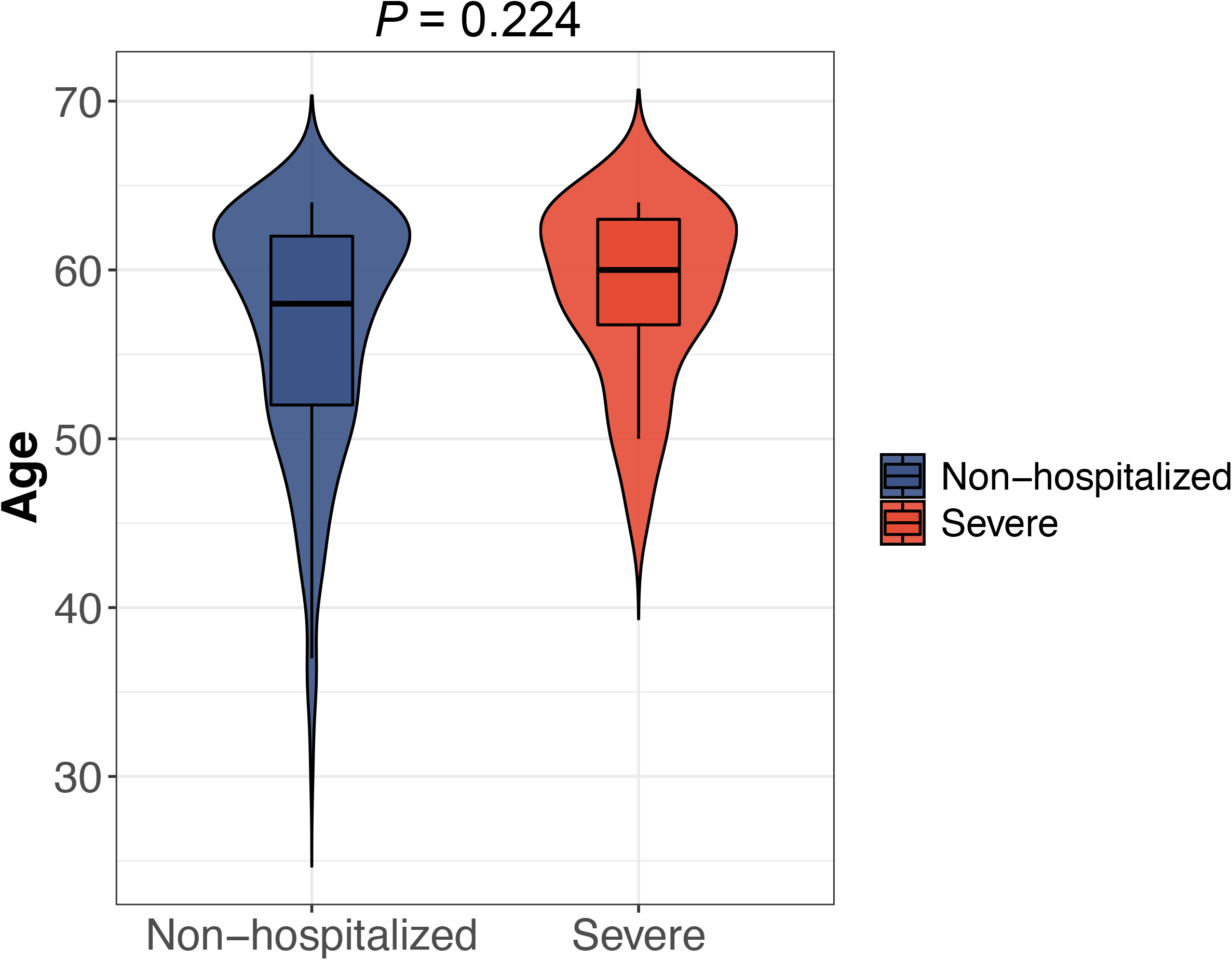
Age distribution of the VA COVID-19 cohort after sample filtering

**Supplementary Figure 8.**
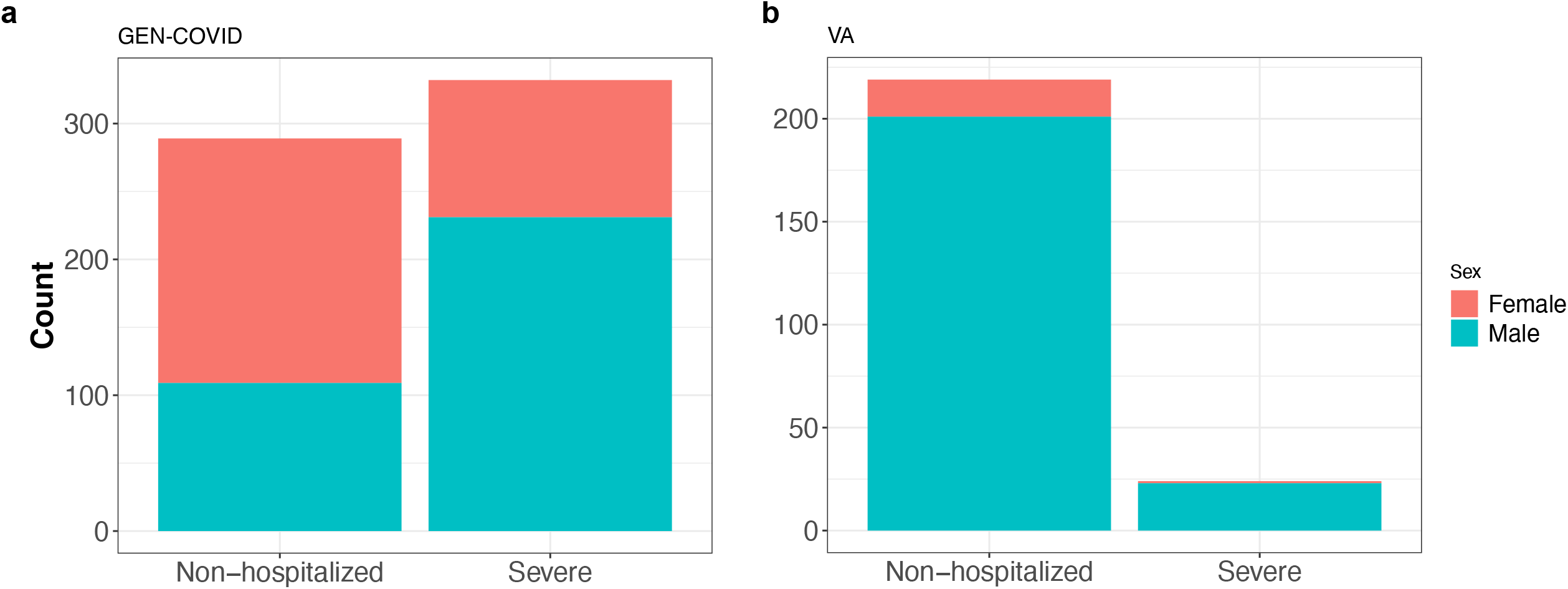
Sex distributions of the GEN-COVID and VA cohorts after sample filtering

**Supplementary Figure 9.**
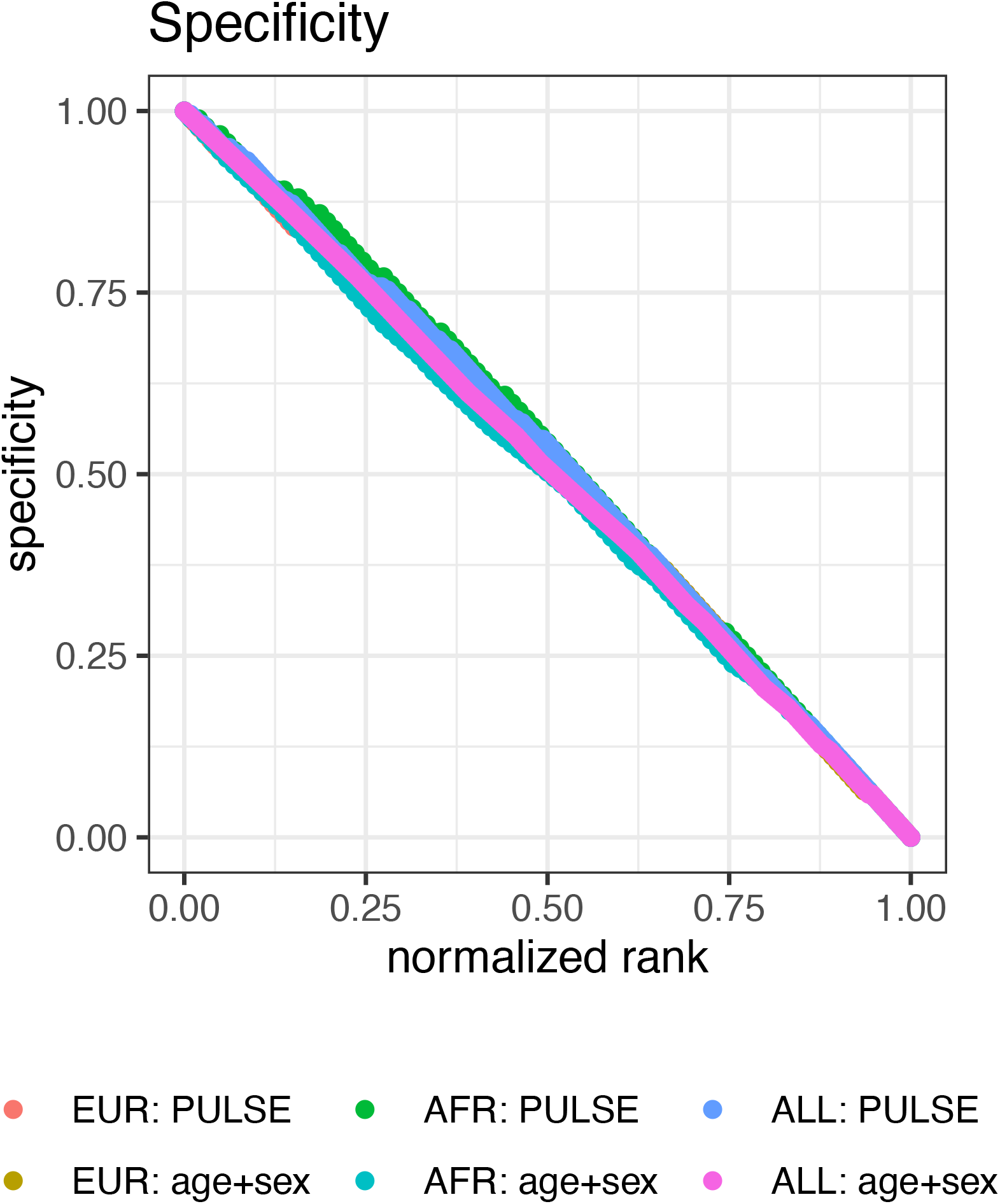
Normalized rank versus specificity of PULSE prediction for the VA COVID-19 cohort

**Supplementary Figure 10.**
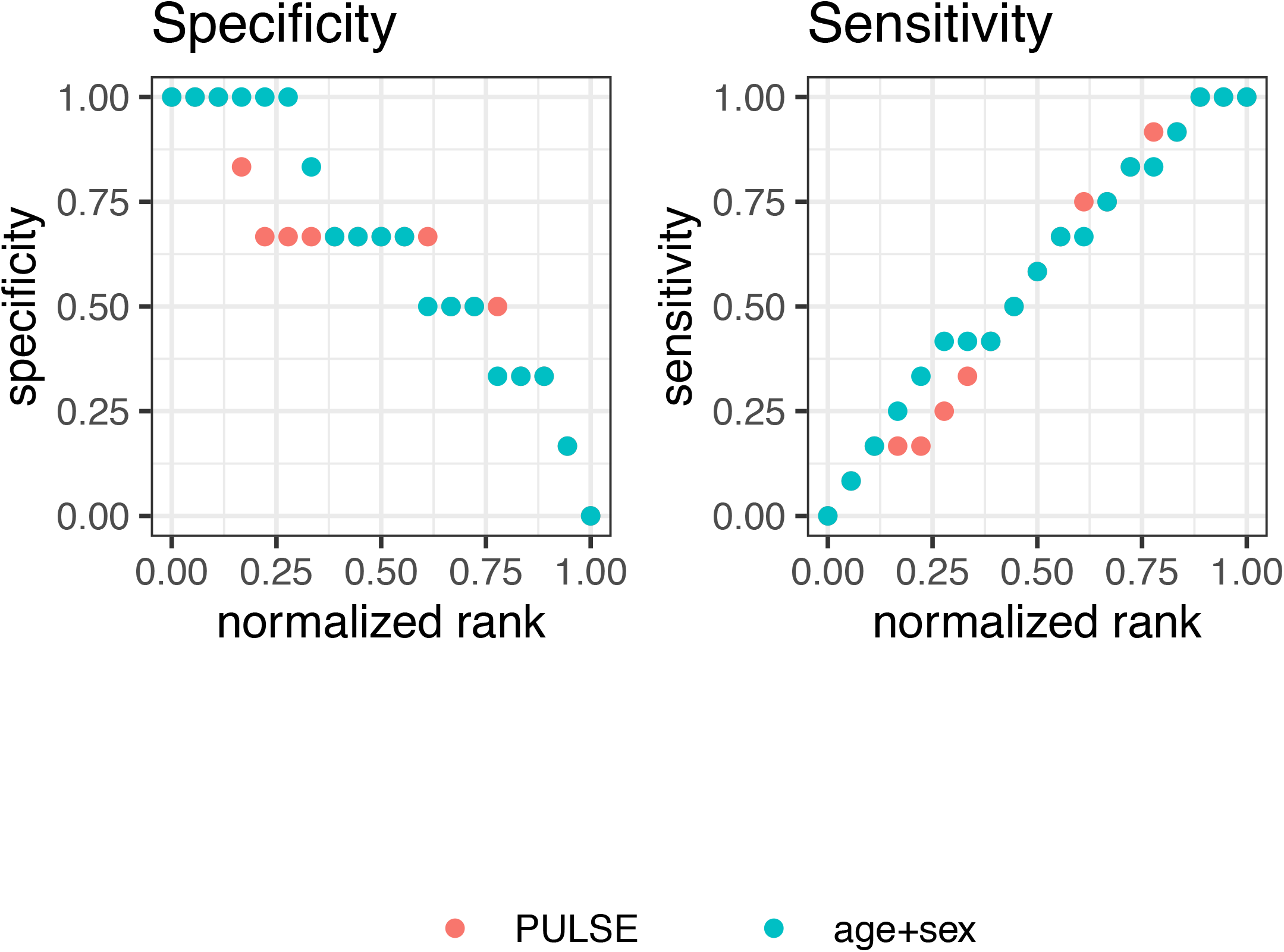
Normalized rank versus specificity and sensitivity of PULSE prediction for GEN-COVID non-EUR samples

**Supplementary Figure 11.**
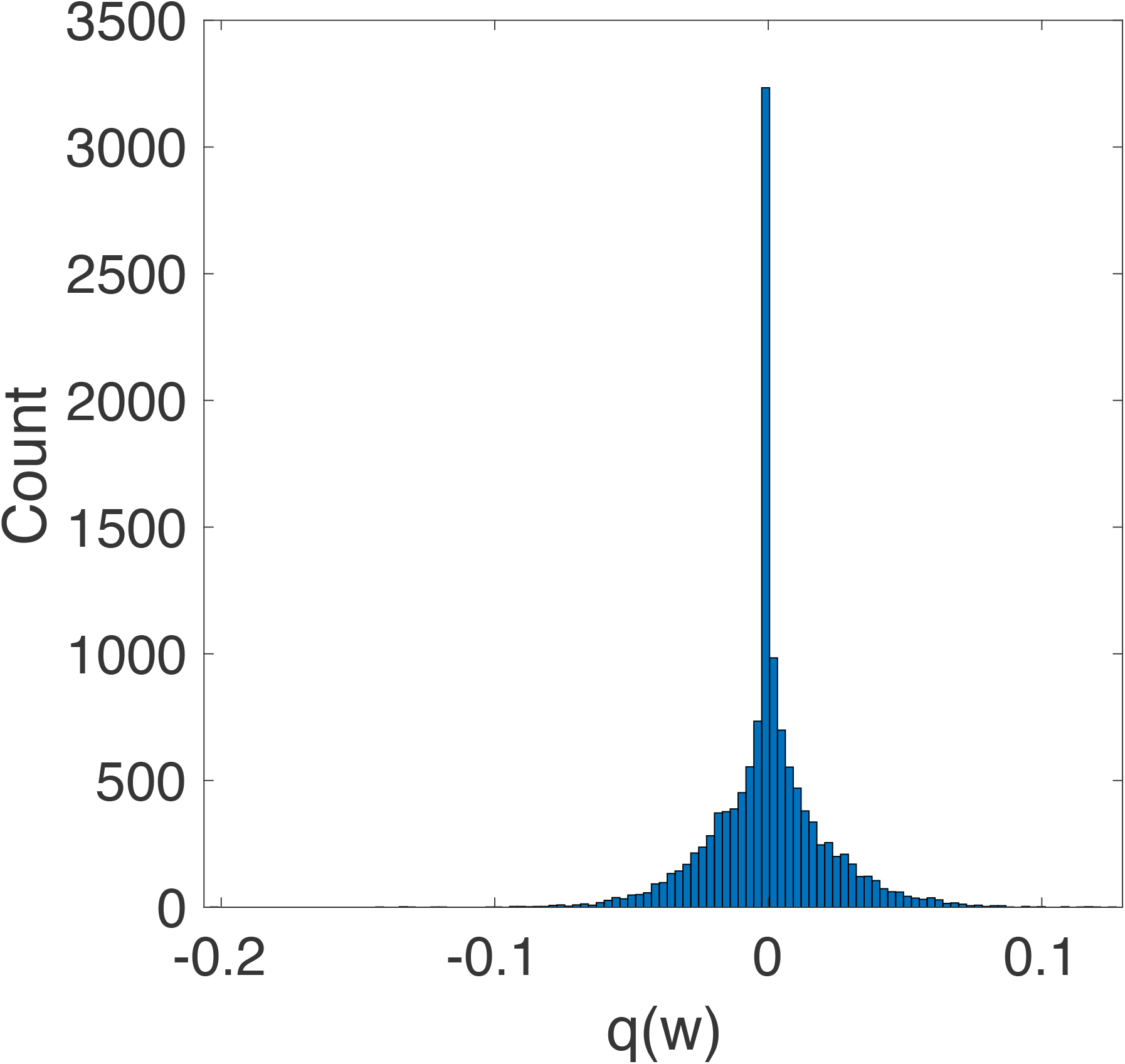
Distribution of gene weights of the PULSE model trained on GEN-COVID EUR samples

### SUPPLEMENTARY TABLES

**Supplementary Table 1**

RefMap COVID-19 regions and genes

**Supplementary Table 2**

Enrichment of disease-associated SNPs in RefMap regions based on the 23andMe study

**Supplementary Table 3**

Partitioned heritability analysis by LDSC for COVID-19 GWAS phenotypes A2, B2, and C2

**Supplementary Table 4**

GO enrichment for RefMap genes per cell type

**Supplementary Table 5**

Pathway enrichment for RefMap genes per cell type

**Supplementary Table 6**

Accession identifiers for ENCODE samples

**Supplementary Table 7**

Variant annotations and their weights learned by PULSE

**Supplementary Table 8**

Clinical characteristics and QC results of 1,339 samples in the GEN-COVID cohort

**Supplementary Table 9**

Clinical characteristics and QC results of 590 samples in the VA COVID-19 cohort

**Supplementary Table 10**

Genes with top 5% weights in the PULSE model

**Supplementary Table 11**

Genes predicted by either RefMap or PULSE to be associated with NK cells

**Supplementary Table 12**

PPI network after network smoothing. Gene identifiers were given in Supplementary Table 13.

**Supplementary Table 13**

Modules detected by the Leiden algorithm and modules significantly enriched with NK-cell COVID-19 genes

**Supplementary Table 14**

GO enrichment for modules enriched with NK-cell COVID-19 genes

**Supplementary Table 15**

Pathway enrichment for modules enriched with NK-cell COVID-19 genes

## Supplementary Notes

Technical details on the PULSE model

## Code availability

The computer codes generated in this study are available from the authors upon request (P.S.T. and M.P.S.).

## Supplementary Notes

### 1. Update rules of variational inference for PULSE

We provide update rules for the local and global variational parameters in `PULSE`.

#### 1.1 Local variational method

As described in the Methods section in our main text, we used the local variational method [1] to handle the sigmoid function in variational inference (VI). Indeed, the sigmoid function involved in the Bernoulli distribution in Eq. 19 in the Methods section can be lower bounded by

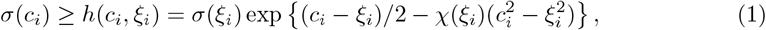

where

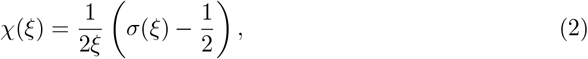

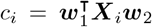 and *ξ*_*i*_ is a local variational parameter introduced to control the bound tightness. Therefore, the log-likelihood of observations is also lower bounded, i.e.,

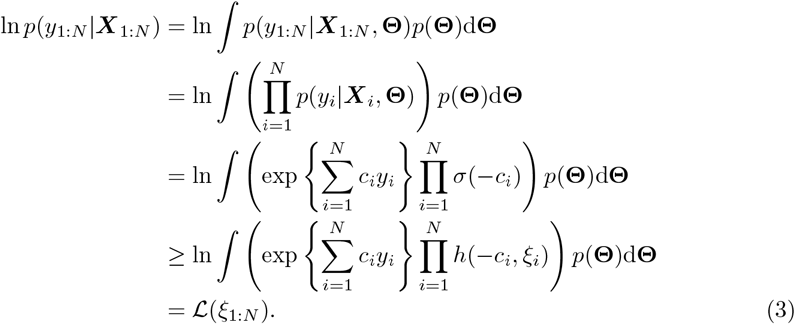

On one hand, we aim to perform variational inference based on the tractability of the lower bound *h*(*c*_*i*_, *ξ*_*i*_), matching the proposal distribution with the true posterior. On the other hand, the variational parameters *ξ*_*i*_’s need to be optimized by maximizing the lower bound ℒ(*ξ*_1:*N*_) of the marginal likelihood, which achieves a better approximation after each update. Therefore, we adopted the variational expectation-maximization (VEM) algorithm that solves both optimization problems simultaneously.

We first note that the “joint distribution”, denoted by 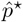, after lower bounding is not a proper density function, but by normalization, the inequality may not hold any more. Indeed, after normalizing, we get

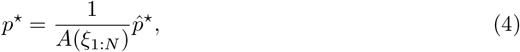

with

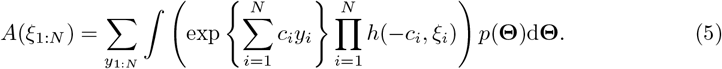

Then, we can rewrite the lower bound ℒ(*ξ*_1:*N*_) as

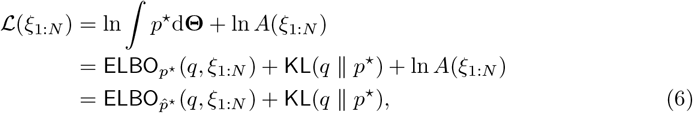

resulting in a similar decomposition of the marginal log-likelihood to that in conventional VI.

As a consequence, we can perform the VEM as follows. (i) In the E-step where the variational parameters ξ_1:N_ are fixed, the standard variational inference is performed to maximize the computationally feasible 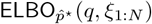 (q, ξ: _1:N_) with respect to q. Here, everything in the mean-field variational inference (MFVI) keeps unchanged except replacing the sigmoid functions in the joint distribution by their lower bounds given by Eq. 1. This computes the approximate distribution best matching the true posterior, i.e., minimizing the KL divergence between q and p^*^ (see the last equation in Eq. 6). After the E-step, we approximately tighten the gap between ℒ(*ξ*_1:*N*_) and the ELBO, and obtain 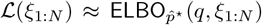 (ii) In the M-step, we fix *q* and maximize the ELBO with respect to *ξ*_1:*N*_, which increases ℒ(*ξ*_1:*N*_) accordingly, as it is obvious that the inequality 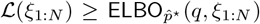 holds. Using VEM, we update *q*’s and *ξ*_*i*_’s iteratively, gradually increasing the log-likelihood lower bound until reaching a local optimum and simultaneously yielding approximate posteriors with performance guarantee.

According to above discussions, we first get the lower bound of the log-likelihood of the conditional distribution over observations, i.e.,

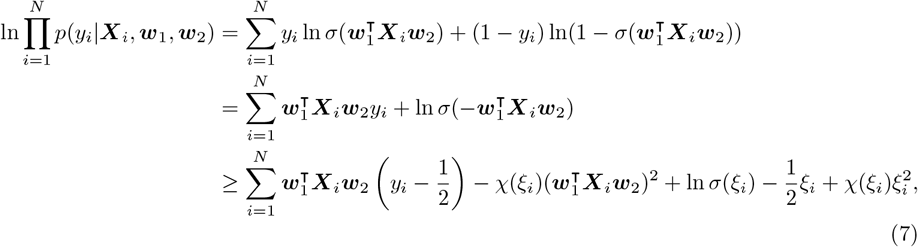

which serves as the basis for the inference of ***w***_1_ and ***w***_2_ Then based on this lower bound and the update principle of MFVI, the logarithm of *q*(***w***_1_) can be calculated as

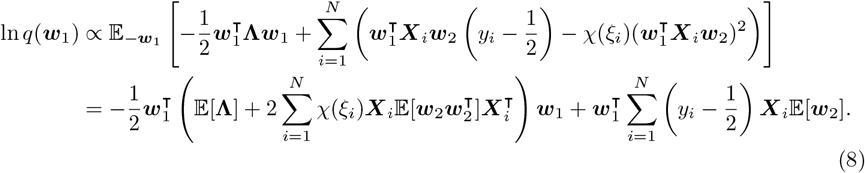

This indicates that *q*(***w***_1_) follows a Gaussian defined as

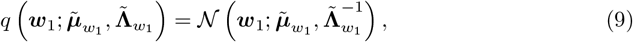

where

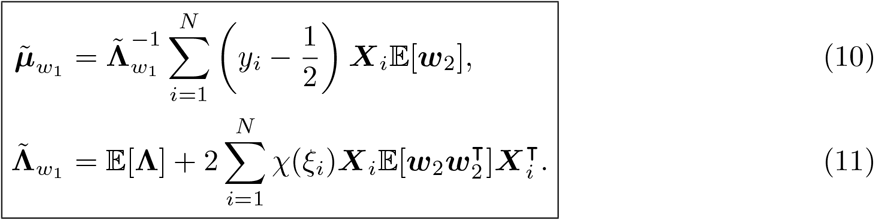

The update rules expressed in Eqs. 10 and 11 are batch based, which is inefficient for large sample size or large feature dimension. We will transform this batch update into stochastic or mini-batch one based on the stochastic variantional inference (SVI), scaling up the inference algorithm to big data. More details are shown in Section 1.3.

#### 1.2 Reparameterization

To perform VI over the spike-and-slab prior defined in Eq. 22 in the Methods section of the main text, we adopted the reparameterization trick introduced in [4]. In particular, as discussed in the Methods section, ***w***_2_ can be reparameterized by two additional variables ***s*** and 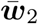 with

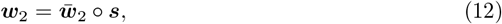

where ∘ means element-wise product. It can be easily shown that the new variable constructed by 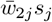 follows the same distribution as *w*_2*j*_. Then we can perform MFVI over 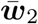 and ***s***. However, the solution derived from a direct application of the fully factorized MFVI will deviate from the true posterior *q*(***w***_/2_) a lot, as the former is unimodal while the latter exponentially multimodal. To solve this problem, we followed [4], in which 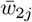 and *s*_*j*_ are bundled together in the factorization. In particular, we assume the proposal distributions factorize as

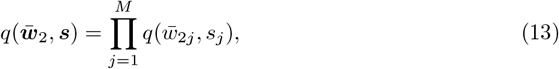

Given the MFVI principle, after substituting *w*_*2j*_ with 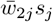 in Eq. 7, we get

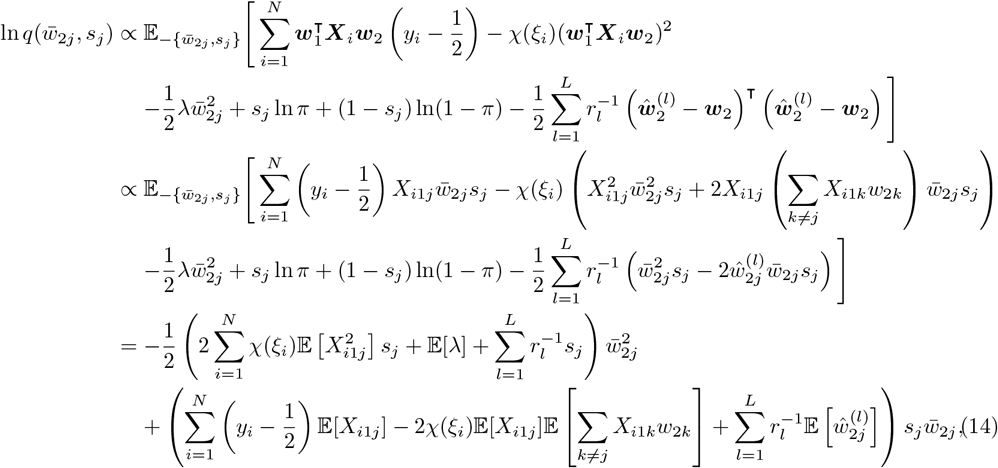

where we define

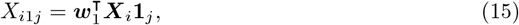

and **1**_*j*_ is a vector with all zeros but the *j*-th element one.

Since 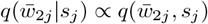, based on Eq. 14, we have

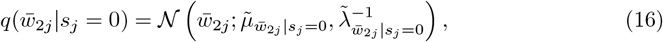

Where

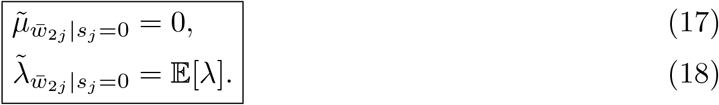

Similarly, 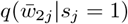 also follows a Gaussian given by

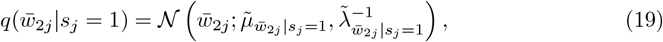

where

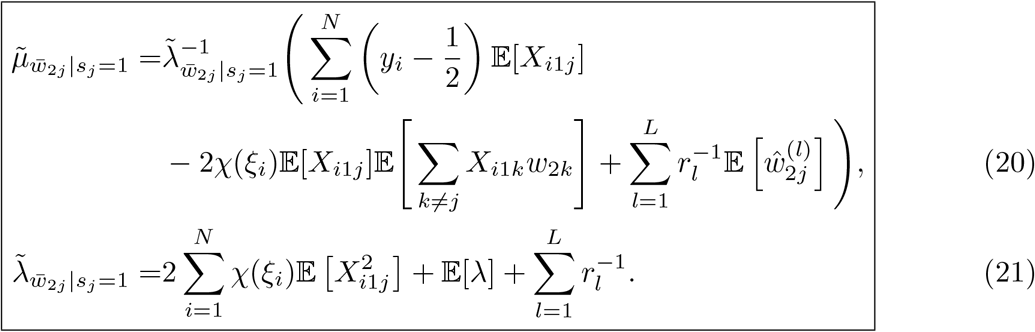

The stochastic updates of Eqs. 20 and 21 are shown in Section 1.3.

To derive *q*(*s*_*j*_), we use the Bayes’ rule given by 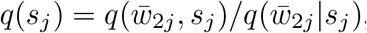, yielding

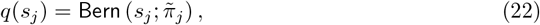

where

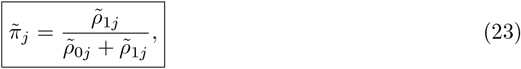

and

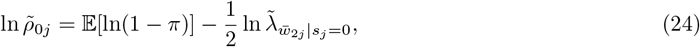

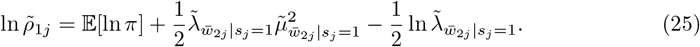

The posterior statistics of *w*_2j_, including the expectation and variance, can be easily calculated based on Eqs. 12, 17, 18, 20 and 21. In particular, the posterior statistics of the marginal 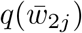 can be derived based on the laws of total expectation and variance, respectively.

#### 1.3 Stochastic variational inference

As discussed in the Methods section in the main text, to scale up the inference algorithm to big data, we adopted SVI proposed in [3]. SVI updates variational parameters by summarizing data points based on stochastic gradient optimization, in which the natural gradient is used to account for measuring similarity between probability distributions. Thanks to the conditional conjugacy introduced in our model, the natural gradient enjoys a simple form without the calculation of the Hessian [3]. Then we can approximate the natural gradient by randomly sampling a single or a mini-batch of samples, greatly reducing the computational complexity per epoch. Here in our inference process, there are two steps where SVI needs to be applied.

(i) For the update of *q*(***w***_1_)whose batch update is given by Eqs. 10 and 11, its stochastic update is given by

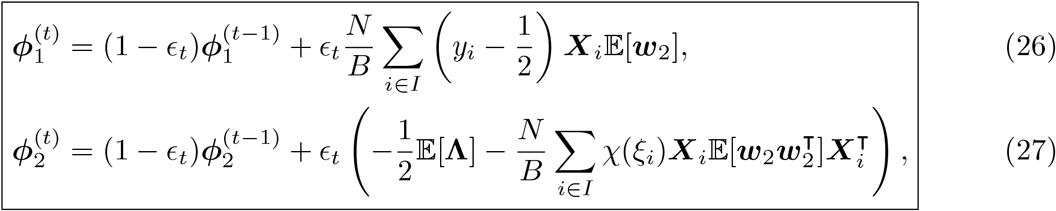

where ***ϕ***_1_ and ***ϕ***_2_ are natural parameters in the exponential family form for multivariate Gaussian, and *I* is a randomly sampled index set from 1: *N* with size *B*. Then the distribuiton parameters in *q*(***w***_1_) can be recovered by

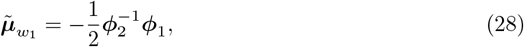

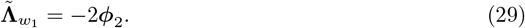

(ii) Similarly, for the update of 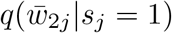, its stochastic version is given by

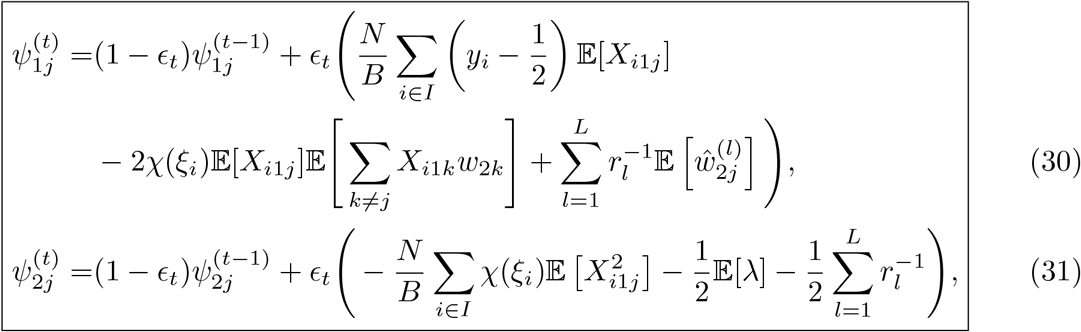

where ***ψ***_1*j*_ and ***ψ***_2*j*_ are natural parameters in the exponential family form of Gaussian. In particular, the parameters in *q*(*w*_1_) can be recovered by

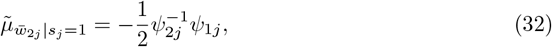

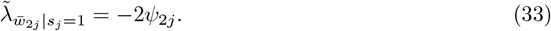

#### 1.4 Update rules for other global variational parameters

For other variational parameters, we perform standard MFVI and have

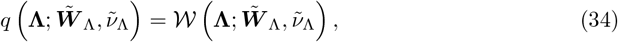

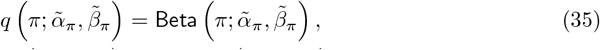

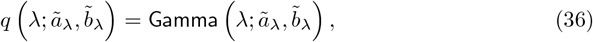

in which

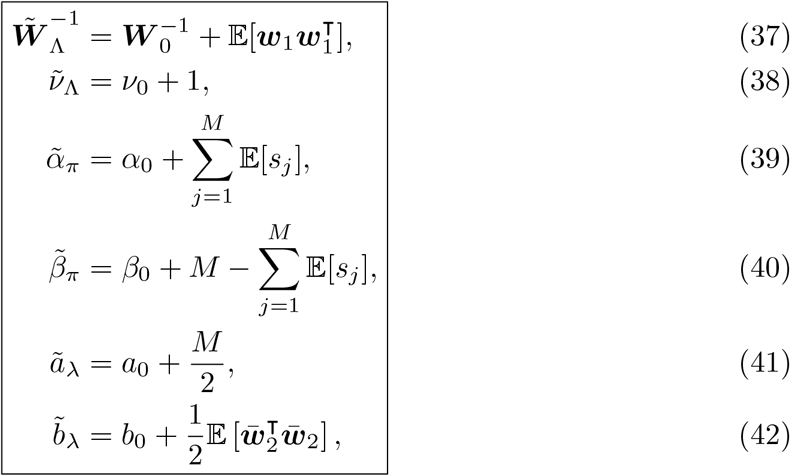

#### 1.5 Update rules for the local variational parameters

In addition to calculating posteriors, we also need to determine the local variational parameters *ξ*_*i*_’s. According to our discussion in Section 1.1, we seek to optimizing *ξ*_*i*_’s by maximizing the lower bound ℒ(*ξ*_1:*N*_) in Eq. 3. This corresponds to the M-step, in which the expected complete-data log-likelihood is maximized, i.e.,

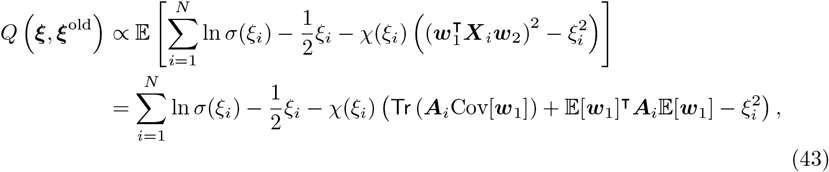

in which

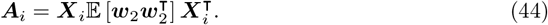

By setting the derivate of Eq. 43 with respect to *ξ*_*i*_ to zero, we get

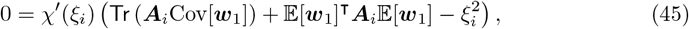

indicating that

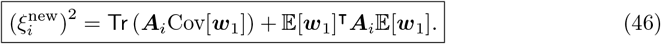

Note that we can force *ξ*_*i*_’s to be nonnegative without loss of generality due to the monotonicity of *χ*(*ξ*_*i*_) when *ξ*_*i*_ ≥ 0.

### 2 Update termination

To terminate the algorithm, we need to monitor the change of ELBO, whose computation is intense and undesirable. In this study, we followed the suggestions proposed in [2], in which we computed the average log predictive for a small held-out dataset to track ELBO evolution. We terminated the updates once the change of average log predictive fell below a threshold, indicating convergence. Here, we set tol = 10^*-*5^ and terminate the algorithm when the proportion of change in ELBO is less than the tolerance. The inference algorithm is summarized in Algorithm 1.

#### Algorithm 1: Stochastic MFVI for PULSE

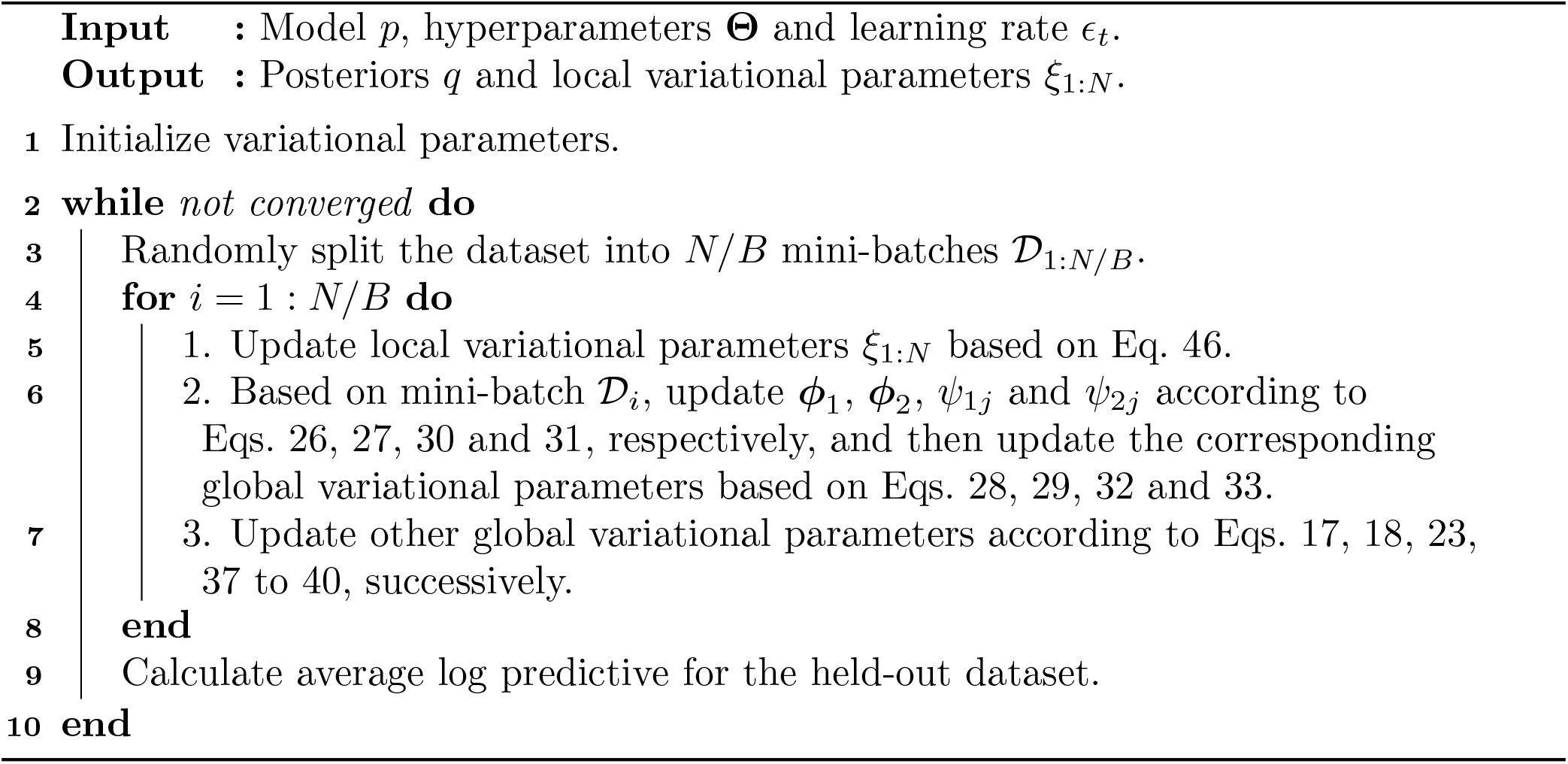

## REFERENCES

1. Dong, E., Du, H. & Gardner, L. An interactive web-based dashboard to track COVID-19 in real time. Lancet Infect. Dis. 20, 533–534 (2020).

2. Shang, Y. et al.. Scoring systems for predicting mortality for severe patients with COVID-19. EClinicalMedicine 24, 100426 (2020).

3. Li, X. et al.. Predictive indicators of severe COVID-19 independent of comorbidities and advanced age: a nested case-control study. Epidemiology & Infection 148, (2020).

4. Initiative, T. C.-19 H. G. & The COVID-19 Host Genetics Initiative. The COVID-19 Host Genetics Initiative, a global initiative to elucidate the role of host genetic factors in susceptibility and severity of the SARS-CoV-2 virus pandemic. European Journal of Human Genetics vol. 28 715–718 (2020).

5. Shelton, J. F. et al.. Trans-ancestry analysis reveals genetic and nongenetic associations with COVID-19 susceptibility and severity. Nature Genetics (2021) doi:10.1038/s41588-021-00854-7.

6. Genomewide Association Study of Severe Covid-19 with Respiratory Failure. N. Engl. J. Med. 383, 1522–1534 (2020).

7. Initiative, C.-19 H. G. & Others. Mapping the human genetic architecture of COVID-19 by worldwide meta-analysis. MedRxiv (2021).

8. Pairo-Castineira, E. et al.. Genetic mechanisms of critical illness in COVID-19. Nature 591, 92–98 (2021).

9. Wang, F. et al.. Initial whole-genome sequencing and analysis of the host genetic contribution to COVID-19 severity and susceptibility. Cell Discovery vol. 6 (2020).

10. Benetti, E. et al.. Clinical and molecular characterization of COVID-19 hospitalized patients. PLoS One 15, e0242534 (2020).

11. Novelli, A. et al.. Analysis of ACE2 genetic variants by direct exome sequencing in 99 SARS-CoV-2 positive patients. (2020).

12. Kosmicki, J. A. et al.. A catalog of associations between rare coding variants and COVID-19 outcomes. medRxiv (2021) doi:10.1101/2020.10.28.20221804.

13. Huang, C. et al.. Clinical features of patients infected with 2019 novel coronavirus in Wuhan, China. Lancet 395, 497–506 (2020).

14. Brodin, P. Immune determinants of COVID-19 disease presentation and severity. Nat. Med. 27, 28–33 (2021).

15. Mehta, P. et al.. COVID-19: consider cytokine storm syndromes and immunosuppression. Lancet 395, 1033–1034 (2020).

16. Mathew, D. et al.. Deep immune profiling of COVID-19 patients reveals distinct immunotypes with therapeutic implications. Science 369, (2020).

17. Sosa-Hernández, V. A. et al.. B Cell Subsets as Severity-Associated Signatures in COVID-19 Patients. Front. Immunol. 11, 611004 (2020).

18. Lucas, C. et al.. Longitudinal analyses reveal immunological misfiring in severe COVID-19. Nature 584, 463–469 (2020).

19. Arunachalam, P. S. et al.. Systems biological assessment of immunity to mild versus severe COVID-19 infection in humans. Science 369, 1210–1220 (2020).

20. Zhang, J.-Y. et al.. Single-cell landscape of immunological responses in patients with COVID-19. Nat. Immunol. 21, 1107–1118 (2020).

21. Stephenson, E. et al.. The cellular immune response to COVID-19 deciphered by single cell multi-omics across three UK centres. medRxiv (2021).

22. Liao, M. et al.. Single-cell landscape of bronchoalveolar immune cells in patients with COVID-19. Nat. Med. 26, 842–844 (2020).

23. Ren, X. et al.. COVID-19 immune features revealed by a large-scale single-cell transcriptome atlas. Cell 184, 1895–1913.e19 (2021).

24. Melms, J. C. et al.. A molecular single-cell lung atlas of lethal COVID-19. Nature (2021) doi:10.1038/s41586-021-03569-1.

25. Delorey, T. M. et al.. COVID-19 tissue atlases reveal SARS-CoV-2 pathology and cellular targets. Nature 1–8 (2021).

26. Miorin, L. et al.. SARS-CoV-2 Orf6 hijacks Nup98 to block STAT nuclear import and antagonize interferon signaling. Proc. Natl. Acad. Sci. U. S. A. 117, 28344–28354 (2020).

27. Blanco-Melo, D. et al.. Imbalanced Host Response to SARS-CoV-2 Drives Development of COVID-19. Cell 181, 1036–1045.e9 (2020).

28. Vietzen, H. et al.. Deletion of the NKG2C receptor encoding KLRC2 gene and HLA-E variants are risk factors for severe COVID-19. Genet. Med. (2021) doi:10.1038/s41436-020-01077-7.

29. Wang, E. Y. et al.. Diverse Functional Autoantibodies in Patients with COVID-19. Nature (2021) doi:10.1038/s41586-021-03631-y.

30. Maucourant, C. et al.. Natural killer cell immunotypes related to COVID-19 disease severity. Sci Immunol 5, (2020).

31. Azzi, Y., Bartash, R., Scalea, J., Loarte-Campos, P. & Akalin, E. COVID-19 and Solid Organ Transplantation: A Review Article. Transplantation 105, 37–55 (2021).

32. Zhang, S., Cooper-Knock, J., Weimer, A. K., Shi, M. & Moll, T. Genome-wide Identification of the Genetic Basis of Amyotrophic Lateral Sclerosis. (2020).

33. Wang, A. et al.. Single-cell multiomic profiling of human lungs reveals cell-type-specific and age-dynamic control of SARS-CoV2 host genes. Elife 9, (2020).

34. Bulik-Sullivan, B. K. et al.. LD Score regression distinguishes confounding from polygenicity in genome-wide association studies. Nat. Genet. 47, 291–295 (2015).

35. Delorey, T. M. et al.. A single-cell and spatial atlas of autopsy tissues reveals pathology and cellular targets of SARS-CoV-2. bioRxiv (2021) doi:10.1101/2021.02.25.430130.

36. Smith, G. D. Mendelian Randomization for Strengthening Causal Inference in Observational Studies. Perspectives on Psychological Science vol. 5 527–545 (2010).

37. Roederer, M. et al.. The genetic architecture of the human immune system: a bioresource for autoimmunity and disease pathogenesis. Cell 161, 387–403 (2015).

38. Raulet, D. H. Roles of the NKG2D immunoreceptor and its ligands. Nat. Rev. Immunol. 3, 781–790 (2003).

39. Travaglini, K. J. et al.. A molecular cell atlas of the human lung from single-cell RNA sequencing. Nature 587, 619–625 (2020).

40. Kuleshov, M. V. et al.. Enrichr: a comprehensive gene set enrichment analysis web server 2016 update. Nucleic Acids Res. 44, W90–7 (2016).

41. Balboa, M. A., Balsinde, J., Aramburu, J., Mollinedo, F. & López-Botet, M. Phospholipase D activation in human natural killer cells through the Kp43 and CD16 surface antigens takes place by different mechanisms. Involvement of the phospholipase D pathway in tumor necrosis factor alpha synthesis. J. Exp. Med. 176, 9–17 (1992).

42. Watzl, C. & Long, E. O. Signal transduction during activation and inhibition of natural killer cells. Curr. Protoc. Immunol. Chapter 11, Unit 11.9B (2010).

43. Mikulak, J., Oriolo, F., Zaghi, E., Di Vito, C. & Mavilio, D. Natural killer cells in HIV-1 infection and therapy. AIDS 31, 2317–2330 (2017).

44. Nuvor, S. V., van der Sande, M., Rowland-Jones, S., Whittle, H. & Jaye, A. Natural Killer Cell Function Is Well Preserved in Asymptomatic Human Immunodeficiency Virus Type 2 (HIV-2) Infection but Similar to That of HIV-1 Infection When CD4 T-Cell Counts Fall. Journal of Virology vol. 80 2529–2538 (2006).

45. Hoffmann, M. et al.. SARS-CoV-2 Cell Entry Depends on ACE2 and TMPRSS2 and Is Blocked by a Clinically Proven Protease Inhibitor. Cell 181, 271–280.e8 (2020).

46. De Biasi, S. et al.. Marked T cell activation, senescence, exhaustion and skewing towards TH17 in patients with COVID-19 pneumonia. Nat. Commun. 11, 3434 (2020).

47. He, L. et al.. Pericyte-specific vascular expression of SARS-CoV-2 receptor ACE2 – implications for microvascular inflammation and hypercoagulopathy in COVID-19. doi:10.1101/2020.05.11.088500.

48. ENCODE Project Consortium et al. Expanded encyclopaedias of DNA elements in the human and mouse genomes. Nature 583, 699–710 (2020).

49. Gel, B. et al.. regioneR: an R/Bioconductor package for the association analysis of genomic regions based on permutation tests. Bioinformatics btv562 (2015) doi:10.1093/bioinformatics/btv562.

50. Lee, S. et al.. Optimal unified approach for rare-variant association testing with application to small-sample case-control whole-exome sequencing studies. Am. J. Hum. Genet. 91, 224–237 (2012).

51. Benetti, E. et al.. ACE2 gene variants may underlie interindividual variability and susceptibility to COVID-19 in the Italian population. Eur. J. Hum. Genet. 28, 1602–1614 (2020).

52. Daga, S. et al.. Employing a systematic approach to biobanking and analyzing clinical and genetic data for advancing COVID-19 research. Eur. J. Hum. Genet. (2021) doi:10.1038/s41431-020-00793-7.

53. Wang, K., Li, M. & Hakonarson, H. ANNOVAR: functional annotation of genetic variants from high-throughput sequencing data. Nucleic Acids Res. 38, e164 (2010).

54. 1000 Genomes Project Consortium et al. A global reference for human genetic variation. Nature 526, 68–74 (2015).

55. Aschard, H. et al.. Combining effects from rare and common genetic variants in an exome-wide association study of sequence data. BMC Proc. 5 Suppl 9, S44 (2011).

56. Pritchard, J. K. Are rare variants responsible for susceptibility to complex diseases? Am. J. Hum. Genet. 69, 124–137 (2001).

57. Pritchard, J. K. & Cox, N. J. The allelic architecture of human disease genes: common disease–common variant… or not? Hum. Mol. Genet. 11, 2417–2423 (2002).

58. Li, J. et al.. Decoding the Genomics of Abdominal Aortic Aneurysm. Cell 174, 1361–1372.e10 (2018).

59. Li, J., Li, X., Zhang, S. & Snyder, M. Gene-Environment Interaction in the Era of Precision Medicine. Cell 177, 38–44 (2019).

60. Szklarczyk, D. et al.. STRING v11: protein-protein association networks with increased coverage, supporting functional discovery in genome-wide experimental datasets. Nucleic Acids Res. 47, D607–D613 (2019).

61. Krishnan, A. et al.. Genome-wide prediction and functional characterization of the genetic basis of autism spectrum disorder. Nat. Neurosci. 19, 1454–1462 (2016).

62. Traag, V. A., Waltman, L. & van Eck, N. J. From Louvain to Leiden: guaranteeing well-connected communities. Sci. Rep. 9, 5233 (2019).

63. Shilo, S., Rossman, H. & Segal, E. Signals of hope: gauging the impact of a rapid national vaccination campaign. Nat. Rev. Immunol. 21, 198–199 (2021).

64. Darby, A. C. & Hiscox, J. A. Covid-19: variants and vaccination. BMJ vol. 372 771 (2021).

65. Petrilli, C. M. et al.. Factors associated with hospital admission and critical illness among 5279 people with coronavirus disease 2019 in New York City: prospective cohort study. BMJ 369, m1966 (2020).

66. Rölle, A. et al.. IL-12–producing monocytes and HLA-E control HCMV-driven NKG2C+ NK cell expansion. J. Clin. Invest. 124, 5305–5316 (2014).

67. Medzhitov, R. & Janeway, C. A., Jr. Decoding the patterns of self and nonself by the innate immune system. Science 296, 298–300 (2002).

68. Hu, W., Wang, G., Huang, D., Sui, M. & Xu, Y. Cancer Immunotherapy Based on Natural Killer Cells: Current Progress and New Opportunities. Front. Immunol. 10, 1205 (2019).

69. Chua, R. L. et al.. COVID-19 severity correlates with airway epithelium–immune cell interactions identified by single-cell analysis. Nat. Biotechnol. 38, 970–979 (2020).

70. Siedner, M. J., Tumarkin, E. & Bogoch, I. I. HIV post-exposure prophylaxis (PEP). BMJ k4928 (2018) doi:10.1136/bmj.k4928.

71. Cheng, S. H. & Higham, N. J. A Modified Cholesky Algorithm Based on a Symmetric Indefinite Factorization. SIAM Journal on Matrix Analysis and Applications vol. 19 1097–1110 (1998).

72. Harva, M. & Kabán, A. Variational learning for rectified factor analysis. Signal Processing vol. 87 509–527 (2007).

73. Blei, D. M., Kucukelbir, A. & McAuliffe, J. D. Variational Inference: A Review for Statisticians. Journal of the American Statistical Association vol. 112 859–877 (2017).

74. Hao, Y. et al.. Integrated analysis of multimodal single-cell data. Cell (2021) doi:10.1016/j.cell.2021.04.048.

75. Quinlan, A. R. & Hall, I. M. BEDTools: a flexible suite of utilities for comparing genomic features. Bioinformatics 26, 841–842 (2010).

76. Finucane, H. K. et al.. Partitioning heritability by functional annotation using genome-wide association summary statistics. Nat. Genet. 47, 1228–1235 (2015).

77. Sun, B. B. et al.. Genomic atlas of the human plasma proteome. Nature 558, 73–79 (2018).

78. Suhre, K. et al.. Connecting genetic risk to disease end points through the human blood plasma proteome. Nat. Commun. 8, 14357 (2017).

79. Choi, K. W. et al.. Assessment of Bidirectional Relationships Between Physical Activity and Depression Among Adults: A 2-Sample Mendelian Randomization Study. JAMA Psychiatry 76, 399–408 (2019).

80. Wootton, R. E. et al.. Evaluation of the causal effects between subjective wellbeing and cardiometabolic health: mendelian randomisation study. BMJ 362, k3788 (2018).

81. Julian, T. H. et al.. Physical exercise is a risk factor for amyotrophic lateral sclerosis: Convergent evidence from mendelian randomisation, transcriptomics and risk genotypes. doi:10.1101/2020.11.24.20238063.

82. Purcell, S. et al.. PLINK: a tool set for whole-genome association and population-based linkage analyses. Am. J. Hum. Genet. 81, 559–575 (2007).

83. Machiela, M. J. & Chanock, S. J. LDlink: a web-based application for exploring population-specific haplotype structure and linking correlated alleles of possible functional variants. Bioinformatics 31, 3555–3557 (2015).

84. Hartwig, F. P., Davies, N. M., Hemani, G. & Davey Smith, G. Two-sample Mendelian randomization: avoiding the downsides of a powerful, widely applicable but potentially fallible technique. Int. J. Epidemiol. 45, 1717–1726 (2016).

85. Burgess, S. & Thompson, S. G. Interpreting findings from Mendelian randomization using the MR-Egger method. Eur. J. Epidemiol. 32, 377–389 (2017).

86. Burgess, S. et al.. Guidelines for performing Mendelian randomization investigations. Wellcome Open Research 4, (2019).

87. Burgess, S., Thompson, S. G. & CRP CHD Genetics Collaboration. Avoiding bias from weak instruments in Mendelian randomization studies. Int. J. Epidemiol. 40, 755–764 (2011).

88. Bowden, J., Hemani, G. & Smith, G. D. Invited Commentary: Detecting Individual and Global Horizontal Pleiotropy in Mendelian Randomization—A Job for the Humble Heterogeneity Statistic? American Journal of Epidemiology (2018) doi:10.1093/aje/kwy185.

89. Verbanck, M., Chen, C.-Y., Neale, B. & Do, R. Detection of widespread horizontal pleiotropy in causal relationships inferred from Mendelian randomization between complex traits and diseases. Nat. Genet. 50, 693–698 (2018).

90. Bowden, J. et al.. Assessing the suitability of summary data for two-sample Mendelian randomization analyses using MR-Egger regression: the role of the I2 statistic. International Journal of Epidemiology dyw220 (2016) doi:10.1093/ije/dyw220.

91. de Leeuw, C. A., Mooij, J. M., Heskes, T. & Posthuma, D. MAGMA: generalized gene-set analysis of GWAS data. PLoS Comput. Biol. 11, e1004219 (2015).

92. Li, H. & Durbin, R. Fast and accurate long-read alignment with Burrows–Wheeler transform. Bioinformatics 26, 589–595 (2010).

93. Poplin, R. et al.. Scaling accurate genetic variant discovery to tens of thousands of samples. bioRxiv 201178 (2018) doi:10.1101/201178.

94. Gaziano, J. M. et al.. Million Veteran Program: A mega-biobank to study genetic influences on health and disease. J. Clin. Epidemiol. 70, 214–223 (2016).

95. Song, R. J. et al.. Phenome-wide association of 1809 phenotypes and COVID-19 disease progression in the Veterans Health Administration Million Veteran Program. PLoS One 16, e0251651 (2021).

96. Regier, A. A. et al.. Functional equivalence of genome sequencing analysis pipelines enables harmonized variant calling across human genetics projects. Nat. Commun. 9, 4038 (2018).

97. Yang, J., Lee, S. H., Goddard, M. E. & Visscher, P. M. GCTA: a tool for genome-wide complex trait analysis. Am. J. Hum. Genet. 88, 76–82 (2011).

98. Alexander, D. H. & Lange, K. Enhancements to the ADMIXTURE algorithm for individual ancestry estimation. BMC Bioinformatics 12, 246 (2011).

99. Liu, X., Wu, C., Li, C. & Boerwinkle, E. dbNSFP v3.0: A One-Stop Database of Functional Predictions and Annotations for Human Nonsynonymous and Splice-Site SNVs. Human Mutation vol. 37 235–241 (2016).

100. Jian, X., Boerwinkle, E. & Liu, X. In silico prediction of splice-altering single nucleotide variants in the human genome. Nucleic Acids Res. 42, 13534–13544 (2014).

101. Lin, H. et al.. RegSNPs-intron: a computational framework for predicting pathogenic impact of intronic single nucleotide variants. Genome Biol. 20, 254 (2019).

102. Wu, B., Guan, W. & Pankow, J. S. On efficient and accurate calculation of significance P-values for sequence kernel association testing of variant set. Ann. Hum. Genet. 80, 123–135 (2016).

103. Mbatchou, J., Barnard, L., Backman, J. & Marcketta, A. Computationally efficient whole genome regression for quantitative and binary traits. bioRxiv (2020).

104. Consortium, G. & GTEx Consortium. Genetic effects on gene expression across human tissues. Nature vol. 550 204–213 (2017).

105. Titsias, M. K. & Lázaro-Gredilla, M. Spike and Slab Variational Inference for Multi-Task and Multiple Kernel Learning. in Advances in Neural Information Processing Systems 24 (eds. Shawe-Taylor, J., Zemel, R. S., Bartlett, P. L., Pereira, F. & Weinberger, K. Q.) 2339–2347 (Curran Associates, Inc., 2011).

106. Hoffman, M. D., Blei, D. M., Wang, C. & Paisley, J. Stochastic variational inference. (2013).

107. Robbins, H. & Monro, S. A Stochastic Approximation Method. Herbert Robbins Selected Papers 102–109 (1985) doi:10.1007/978-1-4612-5110-1_9.

108. Wang, S., Cho, H., Zhai, C., Berger, B. & Peng, J. Exploiting ontology graph for predicting sparsely annotated gene function. Bioinformatics 31, i357–64 (2015).

## References

[1] C. M. Bishop. Pattern Recognition and Machine Learning (Information Science and Statistics). Springer-Verlag, Berlin, Heidelberg, 2006.

[2] D. M. Blei, A. Kucukelbir, and J. D. McAuliffe. Variational inference: A review for statisticians. Journal of the American Statistical Association, 112(518):859–877, 2017.

[3] M. D. Hoffman, D. M. Blei, C. Wang, and J. Paisley. Stochastic variational inference. Journal of Machine Learning Research, 14(4):1303–1347, 2013.

[4] M. K. Titsias and M. Lázaro-Gredilla. Spike and slab variational inference for multi-task and multiple kernel learning. In J. Shawe-Taylor, R. S. Zemel, P. L. Bartlett, F. Pereira, and K. Q. Weinberger, editors, Advances in Neural Information Processing Systems 24, pages 2339–2347. Curran Associates, Inc., 2011.

